# Ten months of temporal variation in the clinical journey of hospitalised patients with COVID-19: an observational cohort

**DOI:** 10.1101/2021.06.01.21258150

**Authors:** ISARIC Clinical Characterisation Group, Matthew Hall

## Abstract

**Background:** There is potentially considerable variation in the nature and duration of the care provided to hospitalised patients during an ongoing infectious disease epidemic or pandemic. Improvements in care and clinician confidence may shorten the time spent as an inpatient, or the need for admission to an intensive care unit (ICU) or high density unit (HDU), while novel treatment modalities may reduce the time course of illness. On the other hand, limited resources at times of high demand may lead to rationing of resources, with less beneficial consequences. Despite little evidence on how the values of such variables change over the course of a crisis (such as the current COVID-19 pandemic), they may nevertheless be used as proxies for disease severity, outcome measures for clinical trials, and to inform planning and logistics. We hypothesise that variation of this kind has been present over the first year of the pandemic.

**Methods and Findings:** We investigate such time trends in an extremely large international cohort of 142,540 patients with symptom onset of, or hospital admission for, COVID-19 during 2020. The variables investigated are time from symptom onset to hospital admission, probability of ICU/HDU admission, time from hospital admission to ICU/HDU admission, case fatality ratio (CFR) and total length of hospital stay. Time from hospital symptom onset to hospital admission showed a rapid decline during the first months of the pandemic followed by peaks during August/September and December. ICU/HDU admission was more frequent from June to August, while there were only modest time trends in time from hospital admission to ICU/HDU. The CFR was lowest from June to August, a trend mostly driven by patients with no ICU/HDU admission. Raw numbers for overall hospital stay showed little overall variation over the time period, but further examination reveals a clear decline in time to discharge for ICU/HDU survivors. The main limitations are that these are predominantly severe COVID-19 cases, and that there are temporal, spatial and demographic biases present in an observational study of this kind.

**Conclusions:** Our results establish that variables of these kinds have limitations when used as outcome measures in a rapidly-evolving situation.

## Introduction

During an epidemic or pandemic of a novel infectious disease, variations in the duration of each stage of a hospitalised patient’s progress from symptom onset, to hospital admission, and hence to outcome are critical for an effective response. Clinicians use these data as a proxy for disease severity, and to provide prognostic information to patients and their families. Policy makers use these data to inform system wide planning for staffing, infrastructure, to predict requirements for consumables (such as personal protective equipment), and to assess performance of the hospital system. And for clinical research, these measures are used as trial outcomes to determine the efficacy of novel treatments.

Often, the extent to which patient journeys vary during an epidemic is not understood. There are changes in clinical practice (World Health Organisation 2021) – clinical understanding of the natural history of diseases improves with time (Docherty et al. 2021), and so too, confidence in safe discharge criteria or alternative models of care (Rojek and Horby 2016), such as remote monitoring (Nunan et al. 2020; Bell et al. 2021). Moreover, the introduction of effective treatments (Rochwerg et al. 2020) and standardisation of care may rapidly reduce the severity or time course of illness (Dennis et al. 2021). However, decisions about whether to admit or escalate care are also dependent on logistic factors such as the availability of resources (e.g. ventilators, intensive care beds, staff) that may be rationed during the peak of a pandemic, but abundant at other phases of an outbreak (Tyrrell et al. 2021; National Institute for Health and Care Excellence 2021; Pagel, Utley, and Ray 2020). There may also be changes in policy to admit patients for indications that are not clinical – such as to facilitate effective quarantine (CMO Messaging 2021) or supervise provision of treatments in clinical trials. We hypothesise that there is significant variation in the patient journey over a pandemic period, and that this variability may limit the way these data can be responsibly used.

In this paper, we assess temporal changes in hospital admission, length of stay, and escalation of care for hospitalized patients with SARS-CoV-2 infection included in the International Severe Acute Respiratory and emerging Infection Consortium (ISARIC) WHO Clinical Characterisation Protocol International cohort (ISARIC Clinical Characterisation Group 2020). This is to our knowledge the largest, prospective international cohort including standardised clinical data, and, as of the time of writing, includes data collected from 26 January 2020 to 7 April 2021 on 483,929 people hospitalised with COVID-19 in 1,609 sites in 54 countries.

We use this dataset to determine whether these variables did indeed change over the course of the SARS-CoV-2 pandemic during 2020, and where there are changes, explore if there are predictable influences that account for this.

## Methods

As previously described (ISARIC Clinical Characterisation Group, Pritchard, and Olliaro 2020), eligible for recruitment were patients with confirmed or suspected COVID-19 infection admitted to an ISARIC partner site and submitted to the ISARIC-hosted REDCap system. Additional data contributed to ISARIC via other mechanisms have not been included due to differences in data structure. The datasets used in analyses in this paper are drawn from a population of all patients with a symptom onset date, or hospital admission date, recorded between March and December 2020 inclusive. Where patients had a recorded hospital admission date before their symptom onset date, this was taken to be a nosocomial infection. The commencement of any patient’s hospital treatment for COVID-19, hereafter “COVID-19 admission date”, was taken to be the later of the symptom onset date and hospital admission date.

We used the complete dataset described above to explore temporal variation in six variables or collections of variables. Not all patients had recorded information for the variables of interest in each question, so, in each case, a subset was analysed. The questions and data subsets were as follows:

**Question 1:** Variation in the time from symptom onset to hospital admission. Patients were excluded if this variable was not available, or if they had nosocomial infection.

**Question 2:** Variation in the proportion of patients being admitted to an ICU or HDU. Patients were excluded if this variable was not available.

**Question 3:** Variation in the time from COVID-19 admission to ICU/HDU admission. Patients were excluded if they were never admitted to an ICU or HDU, or if this variable was otherwise not available.

**Question 4:** Variation in the overall case fatality rate. Patients were excluded if the final outcome of their hospital stay was either not recorded or recorded as something other than “death” or “discharge” (for example, transfer to another facility).

**Question 5:** Variation in the time from COVID-19 admission to death or discharge. (We describe either as an “outcome”.) Exclusions were as in question 4, as well as patients who had a recorded outcome but no recorded outcome date.

**Question 6:** Variation in the status of patients (admitted, ICU/HDU admitted, dead, discharged, or unknown outcome) on a given day after admission. Excluded here were patients whose ICU/HDU status on the day of admission was unknown.

Further filtering was done to a) remove any nonsensical values (such as recorded time of hospital admission after hospital exit), b) remove patients admitted to hospital in 2021 for all questions except 1 (such patients were included when exploring the latter due to right censoring concerns if they were omitted), and c) where a time period was the variable of interest (questions 1, 3, 5 and 6), remove patients with the top 2.5% of values of those variables (as this range included outliers that may have been the result of incorrect data entry).

For all analyses with a single outcome variable, we plotted its mean value against the epidemiological week of symptom onset (question 1) or COVID-19 admission (others), both overall and with respect to various variables of interest (e.g. age group).

For the exploration of patient status by day after COVID-19 admission (question 6), the progress of a patient across the course of their hospital stay was visualised by means of Sankey diagrams. Five states were considered: ward occupancy, ICU/HDU occupancy, the final outcomes of death or discharge, and unknown outcome. We recorded the state of each patient on the day of admission, on every subsequent day, and their final outcome. Where a patient’s exact location (ward or ICU/HDU) in the hospital was not recorded on a given day, their last known location was used. For the figures in this article we present only the data on day of COVID-19 admission (A), three days later (A+3), seven days later (A+7) and final outcome (O+1). We have also developed an interactive version which we hope to make available to the research community as soon as possible.

We used two special categories of symptoms at admission: “common” symptoms (cough, fatigue, fever, and shortness of breath) and gastrointestinal symptoms (abdominal pain, diarrhoea, and vomiting). We introduced variables to the dataset counting the number of each of these categories present for each patient. Missing data was disregarded here, so these represent lower bounds.

### Statistical analysis

Multivariable linear regression was used to investigate factors associated with time from onset of symptoms to hospital admission (question 1), time from COVID-19 admission to ICU/HDU admission (question 3), and time from COVID-19 admission to death or discharge (question 5). In all cases the dependent variable was log-transformed and a pseudocount of 1 added in order to prevent taking logarithms of zero. Multivariable logistic regression was used to investigate factors associated with ICU/HDU admission (question 2) and fatal outcome (question 5). (For a full list of variables used, see supplementary text S1.) For all regression analyses we analysed the presence of comorbidities as covariables. As there was a considerable amount of missing data for each of these, we introduced an “unknown” class to the regression models for these variables rather than exclude patients without values for them entirely. After this modification, every regression analysis was performed as a complete case analysis.

The statistical significance of the variables for month of admission or onset, for age, and for the inclusion of interaction terms used in the regressions for CFR (question 4) and time to outcome (question 5), was assessed using Wald tests.

### Software

All analyses were performed in R 4.0.3 (R Core Team 2013), with packages including the *tidyverse* (Wickham et al. 2019), and *ggalluvial* (Brunson 2020).

## Results

### Patient characteristics

Our complete dataset consisted of 142,540 patients (60,977 female, 81,325 male, 238 unknown sex), median age 70 [IQR 56-82]), admitted at 620 sites in 47 countries. Table 1 shows a summary of baseline characteristics, and more detail, including country of origin and cross tabulation by month of admission, can be found in table S1. Table 2 shows the prevalence of symptoms at admission and comorbidities. 1030 individuals (0.7%) were pregnant women. (For more details, see supplementary table S1.)

**Table 1:** Baseline characteristics of the included patients. *Some patients admitted in early 2021 are included in order to fully represent patients with symptom onset in December 2020.

| Variable | Value | Number of patients | Percent |
| --- | --- | --- | --- |
| **Total** |  | 142540 | 100 |
| **Date of COVID-19 admission**<br>**(month of 2020, or 2021)** | March | 27108 | 19 |
|  | April | 42267 | 29.7 |
|  | May | 12311 | 8.64 |
|  | June | 5342 | 3.75 |
|  | July | 2811 | 1.97 |
|  | August | 2218 | 1.56 |
|  | September | 5265 | 3.69 |
|  | October | 13822 | 9.7 |
|  | November | 15155 | 10.6 |
|  | December | 13205 | 9.26 |
|  | 2021* | 3036 | 2.13 |
| **Sex** | Female | 60977 | 42.8 |
|  | Male | 81325 | 57.1 |
|  | Unknown | 238 | 0.167 |
| **Age group** | 0-19 | 2730 | 1.92 |
|  | 20-39 | 9546 | 6.7 |
|  | 40-59 | 31368 | 22 |
|  | 60-69 | 23431 | 16.4 |
|  | 70-79 | 30541 | 21.4 |
|  | 80+ | 42092 | 29.5 |
|  | Unknown | 2832 | 1.99 |
| **Nosocomial infection** | No | 121910 | 85.5 |
|  | Yes | 11695 | 8.2 |
|  | Unknown | 8935 | 6.27 |

**Table 2:** Prevalence of symptoms at hospital admission and comorbidities. The final column gives the number of times the condition is recorded as present over the number of times its presence or absence is recorded (i.e. the data is non-missing). Designated “common” symptoms are indicated with a (C) and gastrointestinal symptoms with a (G); the number and percentages of patients presenting with combinations of these are separately presented.

|  | Name | % present | *n* present/*n* data recorded |
| --- | --- | --- | --- |
| Symptoms at admission | Cough (C) | 66.8 | 89438/133809 |
|  | Shortness of breath (C) | 64.8 | 92129/142275 |
|  | Fever (C) | 63.4 | 86494/136344 |
|  | Fatigue (C) | 44.9 | 54263/120816 |
|  | Confusion | 24.7 | 31604/127817 |
|  | Vomiting (G) | 19.9 | 25135/126299 |
|  | Myalgia | 18.9 | 21482/113862 |
|  | Diarrhoea (G) | 18.2 | 22953/125804 |
|  | Headache | 12 | 13754/114530 |
|  | Abdominal pain (G) | 11 | 13500/122785 |
|  | Ageusia | 9 | 7103/78674 |
|  | Wheezing | 7.6 | 9011/118028 |
|  | Anosmia | 6.9 | 5550/80040 |
|  | Runny nose | 3.4 | 3745/110937 |
|  | Ulcers | 2.1 | 2306/107859 |
|  | Bleeding | 1.7 | 2123/121852 |
|  | Rash | 1.5 | 1744/116123 |
|  | Seizures | 1.5 | 1821/123380 |
|  | Lymphadenopathy | 0.7 | 783/114739 |
|  | Conjunctivitis | 0.5 | 564/115518 |
|  | Ear pain | 0.5 | 489/97145 |
| Number of recorded “common” symptoms (C) | 0 | 7.6 | 10836/142540 |
|  | 1 | 20.5 | 29257/142540 |
|  | 2 | 26.4 | 37681/142540 |
|  | 3 | 29 | 41359/142540 |
|  | 4 | 16.4 | 23407/142540 |
| Number of recorded gastrointestinal symptoms (G) | 0 | 69.3 | 98714/142540 |
|  | 1 | 20.3 | 28874/142540 |
|  | 2 | 8.5 | 12142/142540 |
|  | 3 | 2 | 2810/142540 |
| Comorbidities | Hypertension | 47.5 | 51415/108319 |
|  | Chronic cardiac disease | 29.5 | 38747/131247 |
|  | Diabetes | 16.8 | 20417/121817 |
|  | Chronic pulmonary disease | 16.4 | 22434/136569 |
|  | Chronic kidney disease | 15.6 | 21199/136148 |
|  | Obesity | 14.5 | 17114/118025 |
|  | Asthma | 13.3 | 18147/136243 |
|  | Dementia | 12.7 | 16538/130084 |
|  | Smoking | 12.7 | 7401/58331 |
|  | Chronic neurological disorder | 11.4 | 15447/135673 |
|  | Rheumatological disorder | 11.2 | 14118/126277 |
|  | Malignant neoplasm | 9.3 | 12548/135418 |
|  | Chronic haematologic disease | 4.1 | 5199/126563 |
|  | Liver disease | 3.4 | 4515/131580 |
|  | Malnutrition | 2.6 | 3125/122243 |
|  | HIV/AIDS | 0.4 | 525/122021 |

### Time from symptom onset to hospital admission (question 1)

After excluding individuals with apparent nosocomial infection, we analysed length of illness before hospitalisation amongst those patients for whom this information was recorded (*n* = 118,980, 83.5%). The median time from symptom onset to admission was 5 days (IQR 1-8). This variable showed a marked decline during March, from a median of 9 days (IQR 5-14) for patients with onset in the week beginning March 1 to 3 (IQR 0-7) in that beginning April 5. Little further variation occurred until late July, when a gradual increase started, which then peaked at a median of 6 (IQR 2-9) for the weeks in late August and early September before a decline to a low of a median 4 (IQR 1-7) days in November; this was followed by another slight increase in December. Times from onset of symptoms to admission were shortest in the oldest and youngest age groups (figure 1b). Patients with a fatal outcome had, generally, shorter times from onset of symptoms until admission compared to survivors (figure 1c).

**Figure 1:**
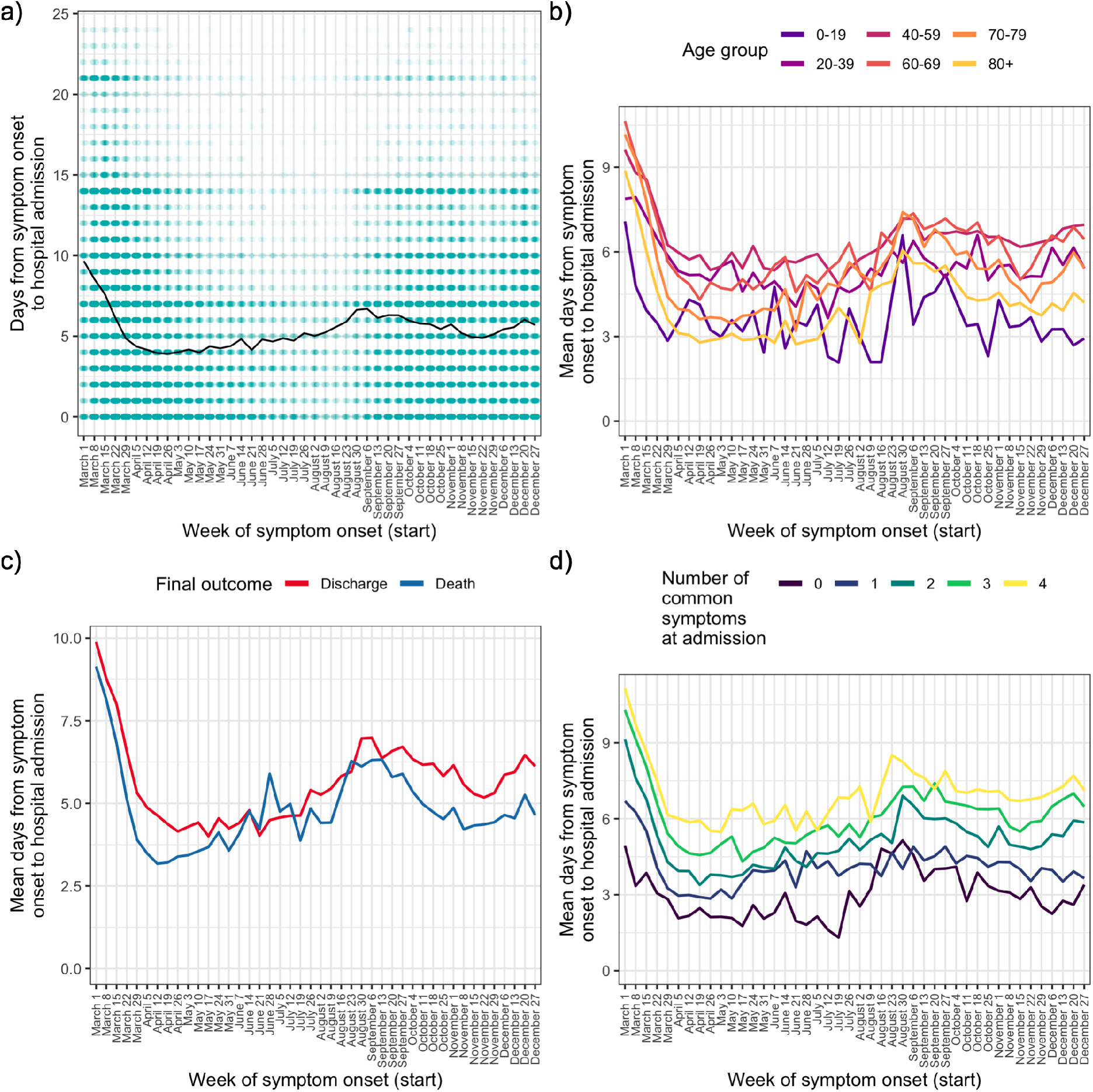
Time from reported symptom onset to hospital admission, by week of reported symptom onset. a) Green cells represent binned patients, with darker colours corresponding to more individuals. The black line represents the mean. b)-d) Mean time to admission plotted by patient characteristics: a) age group, b) final outcome, c) number of the four most common symptoms (cough, fatigue, fever and shortness of breath) present upon admission.

The four most frequent symptoms at admission were cough, fever, shortness of breath, and fatigue; we class these as “common” (see Methods). The number of these that were present increased with time to hospital admission (figure 1d), with the shortest times of all occurring amongst those presenting with none of them. Amongst the 4,636 patients in this analysis presenting with none, the most common other symptoms were confusion (51.6%), vomiting (31.7%), abdominal pain (26%) and diarrhoea (18.9%). Within this group, confusion was the single most common presenting symptom documented in patients over 60, while in younger age groups the most prevalent symptoms were gastrointestinal (see supplementary table S3).

We further explored this question using a multivariable linear regression analysis (supplementary table S4). When compared to April, times from symptom onset to admission were shorter in May and June, and longer in March and from August onwards (all *p*<0.01; see table S4 for confidence intervals). There was very strong evidence of an overall effect of month of onset on time from onset to admission (Wald test *p*<0.001). Patients aged 40-59 showed the longest times to admission when compared to any other age group (all *p*<0.001; see table S4 for CIs). Time from symptom onset to admission was also positively associated with the number of “common” symptoms (25% increase per symptom, 95% CI 24.4%-25.6%), the number of gastrointestinal symptoms (9.8% increase per symptom, 95% CI 9%-10.5%), male sex (4.4% increase, 95% CI 3.4%-5.5%) and discharge as the final outcome (13% increase, 95% CI 11.6%-14.5%).

### ICU/HDU admission and time to ICU/HDU admission (questions 2 and 3)

138,254 (97%) of patients with COVID-19 admission in 2020 had recorded data on whether they ever admitted to an ICU/HDU or not; of these, 29,576 (21.4%) had been admitted at least once. The proportion of individuals with an ICU/HDU admission showed a marked decline over March followed by a renewed peak, and then subsequent decline, in June through August (supplementary figure S1a). The oldest age group (80+) had by far the smallest proportion of ICU/HDU admissions over the whole timeline (5.4%, compared with, for example, 33.8% in the age-group 60-69). In a multivariable logistic regression model (supplementary table S5), the following patterns were observed: there were higher odds of ICU/HDU admission during all months except May, when compared to April (all *p*<0.01, see table S5 for CIs); those aged 80+ had lower odds of ICU/HDU admission (OR 0.12 for admission when compared to the 40-59 age group, 95% CI 0.11-0.13). Males were more likely to be admitted to ICU/HDU (OR 1.53, 95% CI 1.47-1.59). Patients who died had greatly increased odds of having been previously admitted (OR 5.38, 95% CI 5.13-5.65). Compared to those with symptom onset less than a week before hospital admission, patients with nosocomial infections had lower odds of being admitted to ICU/HDU (OR 0.68, 95% CI 0.62-0.74), whereas those with longer times to hospital admission had increased odds (OR 1.36 for 7-13 days, 95% CI 1.31-1.42, 1.28 for 14 or more days, 95% CI 1.2-1.37). An overall effect of month of COVID admission on odds of ICU/HDU admission was highly significant (Wald test *p*<0.0001). Increasing numbers of gastrointestinal symptoms (abdominal pain, vomiting, diarrhoea) were associated with slightly lower odds of admission (OR 0.94 per symptom, 95% CI 0.91-0.96). Comorbidities associated with higher odds of admission were HIV/AIDS (OR 1.46, 95% CI 1.14-1.88), hypertension (OR 1.26, 95% CI 1.2-1.32) and obesity (OR 1.74, 95% CI 1.65-1.83), whereas a wide variety of serious or chronic medical conditions were associated with lower odds (see Table S5), as was smoking (OR 0.82, 95% CI 0.85-0.9). The most extreme fitted odds ratio for a comorbidity with a positive association was 1.74 for obesity, while that for an inverse association was 0.21 for dementia.

We next analysed time from COVID-19 admission in 2020 to first ICU/HDU admission. Excluding those with missing data on this variable, this dataset comprised 26,687 individuals with a maximum time to ICU/HDU of thirteen days. The median time to ICU/HDU was 1 day (IQR 0-2). Raw time trends in this variable were modest (supplementary figure S1b).

Multivariable linear regression (supplementary table S6) nevertheless did show evidence for an overall association with month of COVID-19 admission (Wald test *p*<0.001), with, when compared to April, evidence for longer times to ICU/HDU in March, October and December (all *p*<0.001; see Table S6). Time to ICU/HDU also showed a general increase with age (Wald test for overall association *p*<0.001). There was no evidence of an association with final outcome (death or discharge) or with sex. Time to ICU/HDU admission increased with the number of GI symptoms (9% increase per symptom, 95% CI 7.5%-10.5%). Compared to patients admitted to hospital within a week of symptom onset, those with a nosocomial infection had a 66% increase in time to ICU/HDU (95% CI 54.4%-78.4%) while those with longer times to admission had shorter times to ICU/HDU (9.2% decrease for 7-13 days, 95% CI 7.1%-11.1%, 14.9% decrease for 14 or more days, 95% CI 11.8%-17.8%). Comorbidities associated with longer time to ICU/HDU were asthma (7% increase, 95% CI 3.8%-10.2%), chronic haematological disease (20.6% increase, 95% CI 12.8%-28.9%), and chronic kidney disease (9.8% increase, 95% CI 5.9%-13.9%). In contrast, obesity (7.5% decrease, 95% CI 5.1%-9.9%), diabetes (3.5% decrease, 95% CI 0.7%-6.1%) and smoking (4.9% decrease, 95% CI 0.1%-9.5%) were associated with shorter time to ICU. There was also evidence of a longer time to ICU/HDU amongst pregnant patients (17.5%, 95% CI 3.4%-33.5%).

### Case fatality rate and time from COVID-19 admission to outcome (questions 4 and 5)

We next analysed the final outcome of death or discharge, and the total time from hospital admission to one of those outcomes, in a set of 113,797 patients admitted during 2020 with one of those outcomes recorded (81.8% of the total). The raw case fatality rate (CFR) was 0.3. The median time to death was 8 days (IQR 4-14) and to discharge 7 (IQR 4-14). Over the entire ten-month period of interest (supplementary figure S4), peak CFR was 0.35 in the week beginning 8 March. There was a decline over the spring to a low of 0.17 in the week beginning 12 July, but this trend subsequently reversed and reached 0.32 by mid-December. At the same time, the mean time from admission to outcome in this whole population showed very little change following a dramatic decline during March, from 16 days in the week beginning 1 March to 10 at the start of April (supplementary figure S3). These overall patterns, however, mask substantial variation based on ICU/HDU admission and, in the latter case, outcome (figure 2). The trend in CFR is largely driven by patients who were not admitted to an ICU or HDU. The most consistent decline in time to outcome was observed in ICU/HDU admissions who survived (a decline in the mean of 7.6 days between the first and last weeks studied, figure 2B, bottom left) while survivors with no ICU/HDU admission showed, as with the overall trend, little change after March (bottom right). Variation in time to death appeared very modest amongst patients with an ICU/HDU admission (top left), while there was a distinct peak around August and September in those without (top right). When age is also considered (supplementary figures S5 and S6) a notable additional pattern is the clear correlation of time to discharge and age in surviving non-ICU/HDU patients, which is much less obvious, if present at all, in patients with an ICU/HDU admission.

**Figure 2:**
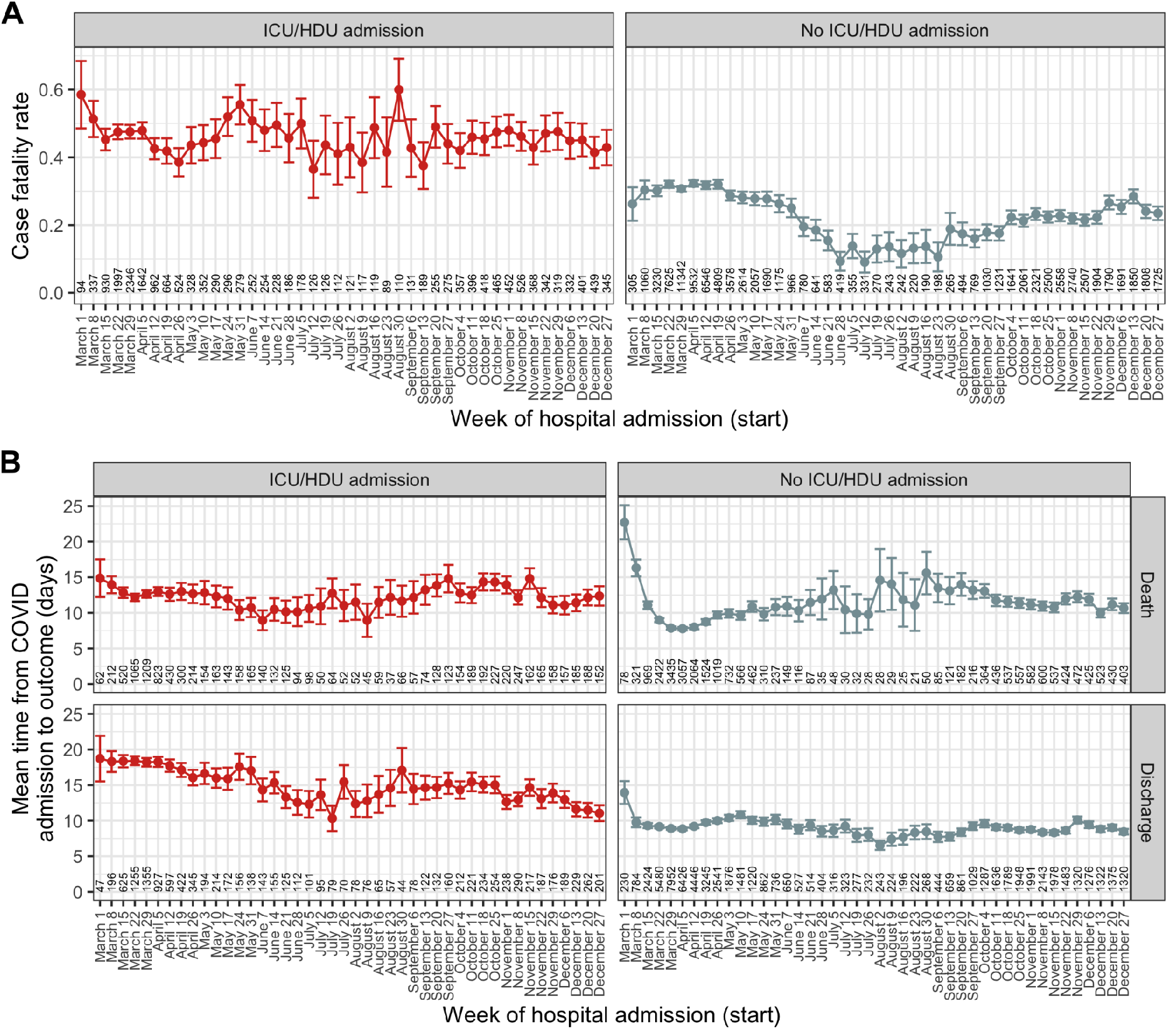
A) Time trends in case fatality ratio, by recorded ICU/HDU admission. B) Time trends from COVID-19 admission to the outcome of death or discharge, further faceted by ICU/HDU admission. Points are mean values, and error bars represent the 95% confidence interval for the mean. Numbers along the x-axis indicate the numbers of patients involved in each category.

The full results of the three multivariable regression analyses can be seen in table 4. There was strong evidence of an association of month of COVID-19 admission with all three variables (Wald test *p*<0.001 in all cases).

**Table 4:**
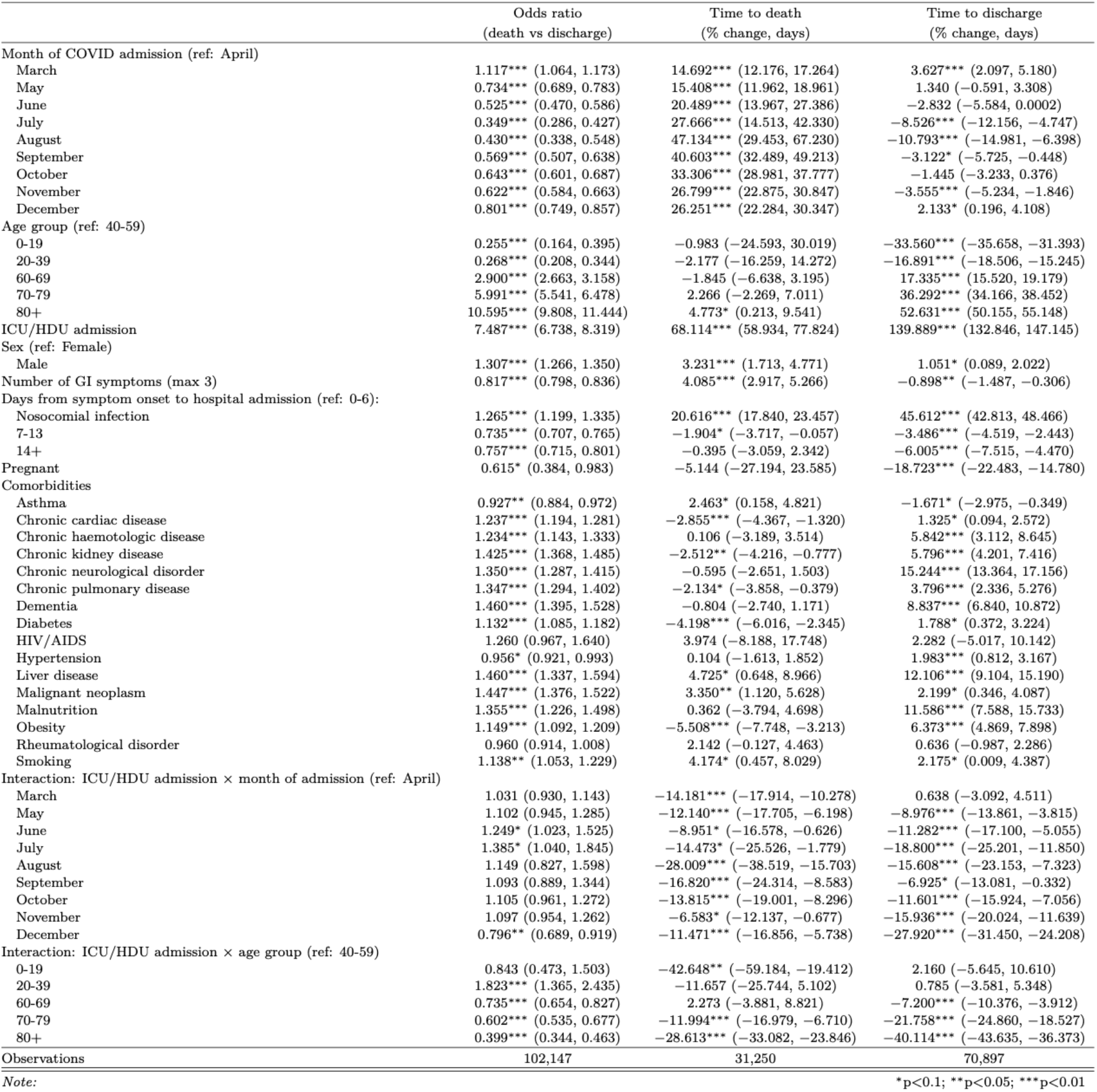
Combined results of logistic regression analysis identifying predictors of death as an outcome, and two linear regression analyses identifying correlates of time to death and time to discharge. For brevity, the country variable, as well as the “unknown” class for each comorbidity (representing patients with missing data for that condition) are omitted here; see table S7 for a version with them included.

Amongst non-ICU/HDU patients, the month with the greatest odds of death was March (OR 1.12 compared to April, 95% CI 1.06-1.17), while that with the smallest was July (OR 0.35, 95% CI 0.29-0.43). April (the reference category) was the month with the shortest time to death, while August had the longest (47.1% increase, 95% CI 29.5%-67.2%). Variation in time to discharge was more modest; the month with the largest value of this variable was March (3.6% increase, 95% CI 2.1%-5.2%), and that with the shortest was August (10.8% decrease, 95% CI 6.4%-15%).

ICU/HDU admission was associated with a 7.5-fold higher odds of death (95% CI 6.74-7.83), a 68.1% increase in time to death (95% CI 58.9%-77.8%) and a 149.9% increase in time to discharge (95% CI 132.8%-147.1%). As a result of the patterns observed in figure 2, we also fitted interaction terms of month of COVID-19 admission with IDU/HCU admission. Their inclusion was consistently statistically significant (Wald test *p*<0.001 in all cases), although for odds of death this ceases to be true when December is removed (*p*=0.24). Hence, the overall increased odds of death amongst ICU/HDU patients was significantly mitigated in December (combined OR 5.96 vs non-ICU/HDU admissions in December, 95% CI 5.13-6.92). There was no evidence that ICU/HDU patients admitted in March and May had a longer time to death than April, but the estimates for all other months were significantly longer, with the peak in November (18.5% increase, 95% CI 13.1%-24.1%). The longest times to discharge were in March (4.3% increase vs April, 95% CI 1.3%-7.3%) and the shortest in December (26.4% decrease, 95% CI 22.8%-29.8%).

Increasing age was associated with monotonic increases in odds of death and time to discharge, with and without ICU/HDU admission. Time to death showed little evidence of variation by age in non-ICU/HDU patients except for marginal evidence for an increase in the oldest age group (4.7% increase vs 40-59, 95% CI 0.2%-9.5%). In ICU/HDU patients, however, where an interaction term was again fitted, the shortest times to death were recorded in both the youngest (43.1% decrease, 95% CI 30.4%-53.6%) and oldest (25.2% decrease, 95% CI 22.2%-28.1%) groups; longest times to death were in middle-aged adults (40-69). (Note that all recorded comorbidities were controlled for here.) Male sex was associated with higher odds of death (OR 1.31, 95% CI 1.27-1.435, and small increases in time to both death (3.2% increase, 95% CI 1.7%-4.8%) and discharge (1.1% increase, 95% CI 0.1%-2%). Nosocomial infection was also associated with higher odds of death (OR 1.27, 95% CI 1.2-1.34) and large increases in time to death (20.6% increase, 17.8%-23.5%) and discharge (45.6% increase, 95% CI 42.8%-48.5%). Patients admitted more than a week from symptom onset had lower odds of death, and shorter stays in hospital, regardless of outcome (see table 4). Where associations with comorbidities were detected, the majority were in the direction of poorer outcomes (increased CFR, decreased time to death, and increased time to discharge), with a few exceptions. Most notably, asthma was associated with lower odds of death (OR 0.92, 95% CI 0.88-0.97), longer times to death (2.5% increase, CI 0.2%-4.8%) and shorter times to discharge (1.7% decrease, 95% CI 0.3%-3%).

### Status by days since admission

Figure 3 displays Sankey diagrams reflecting the location of patients within hospital (ward or ICU) or their final status (dead, discharged, or unknown) on the day of COVID-19 admission (A), three days later (A+3), seven days later (A+7) or, to represent the final status only, one day after the last day in hospital (O+1). The plot is facetted by age group and month of COVID-19 admission. For simplicity, only four months (March, May, June and September) appear in the main figure, but see Supplementary Figure S2 for all months, featuring a total of 129,044 patients (90.5%).

**Figure 3:**
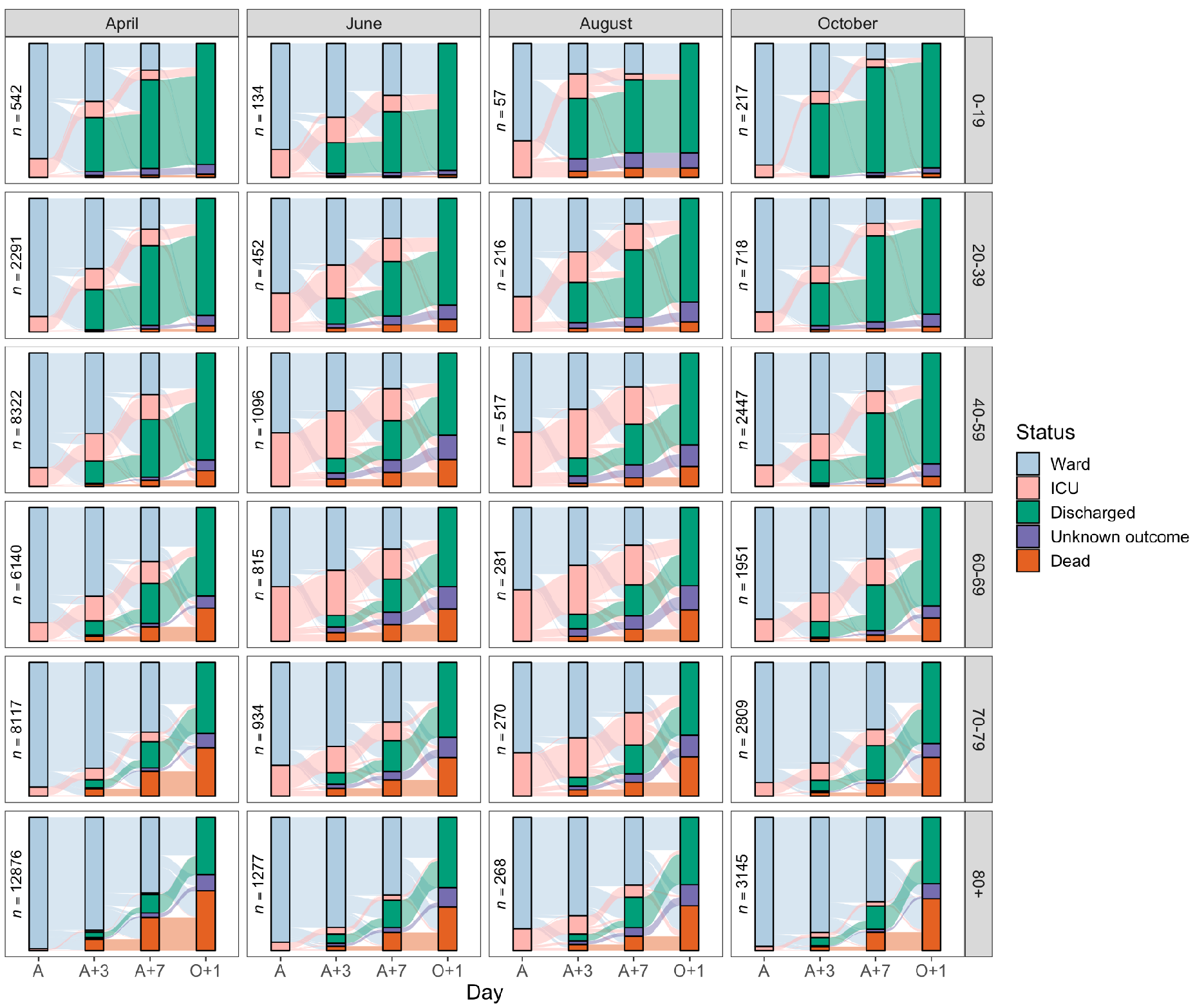
Sankey diagrams depicting the progress through the inpatient journey for patients with COVID-19 admission in April, June, August and October 2020, and subdivided by age. Bars are presented for the day of admission (A), three and seven days later (A+3 and A+7), and the day after final outcome (O+1).

## Discussion

To the best of our knowledge and at the time of publication, this is the largest international cohort of COVID-19 patients in the world. Considerable temporal variations in the events preceding and during hospitalization for patients with confirmed COVID-19 were observed during the period March to November/December 2020. We specifically looked at length of illness before admission, probability of ICU/HDU admission, time to ICU/HDU admission for those so admitted, case fatality rate, and duration of admission overall.

These results highlight key findings with practical implications for case management, resource allocation, performance benchmarking, and reporting of outcomes in research, and point to the fact that patient’s journeys vary over time and must be interpreted with the background of transmission intensity, policy, and practice where cases occur. Therefore, static ‘snapshots’ of the situation at any one time may lead to misguided practice and management if not regularly monitored and approaches adapted accordingly.

In a recent preprint (Kirwan et al. 2021), Kirwan et al analysed temporal variation on time from hospital admission to death, discharge or ICU/HDU admission amongst a smaller cohort of UK patients. We confirm many of the trends that they identified, including the lower CFRs over the summer and the increased odds of ICU/HDU admission in middle-aged age groups. They did not, however, detect the increase in the proportion of patients with an ICU/HDU admission during the summer, or the decline in time to discharge amongst non-ICU/HDU patients over the entire time period. As there were many fewer hospitals included in that study than in ours (31 versus 620) this may be suggestive of variation in available ICU/HDU capacity and usage amongst participating sites in the two studies.

### Prior to admission

Across all age groups, the length of illness before seeking hospital care was longest in July and August when case numbers were lower, and shortest at the extremities of the age distribution and for females. These variations may partially be explained by variations in health-care-seeking behaviour by different demographics, and by differences in clinical progression of disease for different groups. For example, patients who died had consistently shorter duration of illness before hospital admission, possibly reflecting the fact that more serious cases evolve more rapidly and those affected seek care earlier – patients admitted after experiencing symptoms for longer than 1 week were less likely to die. The peak in time to admission during late summer and early autumn in the Northern Hemisphere may reflect delayed presentation following return from holiday, particularly given the high proportion of UK patients in this dataset and known viral importations to the UK from continental Europe around that time (Hodcroft et al. 2020).

At the same time, when considering the four most frequent symptoms at admission (fever, short of breath, cough or fatigue), more symptoms were associated with a longer period between onset of symptoms and admission – and this was consistently so across the entire period under observation. This could be ascribed at least in part to variations in individual behaviour; some patients may present to hospital with a single symptom while others may wait a longer period until several have emerged. In addition, these phenomena could also partially be attributed to how case definitions are applied by physicians, or to the patient’s own perceptions, or to those of their families. Some presentations are likely to be more alarming to the latter two groups than others; for example, individuals with none of the four symptoms described above were admitted fastest of all and, amongst these, confusion was the most prevalent other symptom.

### During hospitalization

Treating variables such as final outcome, ICU/HDU admission, or length of stay, as variables that remain static throughout an evolving epidemic is problematic, as demonstrated by our analyses. To give three examples: first, the case fatality rate showed an overall decline from 0.35 for cases admitted in March to 0.21 in July, followed by a renewed increase to 0.29 in December (see supplementary figure S3). Second, the data underlying the alluvial plots (figure 3) allow us to determine that the proportion of patients discharged within a week of admission rose from 0.24 in March to a peak of 0.34 in September. Third, the proportion of still-admitted patients occupying an ICU/HDU bed showed considerable variation: for example, at day 3 this went from 0.19 in March to 0.13 in April, then rose to a peak of 0.38 in August before declining again, reaching a low of 0.15 in November. Variations in clinical care, the influence of treatments, and changes in available bed capacity are all likely to account for many of these differences. In older patients, the availability of social care space is another important variable.

Patients older than 80 had odds of being admitted to ICU/HDU over eight times smaller than those in the 40-59 category, which may reflect prognosis and the expected benefits of ICU/HDU admission, as well as patient preferences. These decreased odds are also reflective of the temporality of the data. March and April represent our data’s highest volume, which might reflect hospital capacity and the necessity for ICU/HDU prioritization. For the patients who were admitted to ICU, there was no clear trend in the time from hospital admission to transfer to ICU/HDU after March. Length of illness before admission to the hospital and young age were associated with a shorter time from hospital admission to ICU/HDU (for example, a 9.2% decrease for those waiting 7-13 days from onset compared to those waiting less than a week, and 32% decrease amongst under-20s compared to the 40-59 age group), while a smaller proportion of older patients are escalated to ICU/HDU (OR 0.51 for ICU/HDU admission in the 70-79 age group) and after a longer time spent in the ward (a 4.2% increase in the same age group).

### Outcome

As mentioned above, in patients with an outcome of death or discharge, CFRs decreased from 0.35 in March to 0.21 by mid-2020 to increase again to 0.29 in December, mostly following the waves in the pandemic and therefore the number of admissions. System capacity may be an important predictor of patient outcome and may supersede other factors such as increasing case management skills and the influence of new therapies. This also warns against using outcome data that are not adequately controlled to assess efficacy and safety of treatments or other interventions, as effects may rather reflect capacity of a system to provide high-quality care.

We found that shorter time to death is associated with female sex, lack of ICU/HDU admission, and, amongst ICU/HDU patients, the extremes of age. Shorter time to discharge is also associated with female sex and lack of ICU/HDU admission, and this variable increases monotonically with age.

Cautionary notes in interpreting these findings. First, the dataset analysed is made of patients on the more severe end of the spectrum of disease compared to cases occurring in the community. Second, about half of these patients were hospitalized in just two months (March-April), were predominantly from the UK, and about half were over 70 years old. These demographics explain the CFR and the large proportion of patients presenting with age-related comorbidities – nearly half have hypertension, one-in-five chronic cardiac disease, and one-in-six diabetes. Third, there are inherent limitations of observational data, however large the dataset. Fourth, some variables are based on patient self-report which can be inexact; for example it can be clearly seen in figure 1a that multiples of seven reported days from symptom onset to admission are overrepresented, suggesting reports in units of weeks. Fifth, one should refrain from overinterpreting data: some of the changes observed reflect adjustments in practice and logistics, combined pressure on health systems, more than actual effects of interventions. Sixth, resuscitation status and suitability for intensive care admission was not collected in our cohort.

### Implications of findings

Often, in high-income countries, patient outcomes are seen through the lens of individualized treatment provided at the clinician patient interface. This paper demonstrates that outbreak epidemiology has an important influence on patient outcomes – the patient journey from likelihood of admission, through to disposition and length of stay in hospital, and overall outcome, change over the course of a pandemic. There are various explanations for variability – systems may at times be overwhelmed and unable to provide the usual quality of care to their patients; patient behaviour may change depending on perceptions of the status of the outbreak and the performance of the healthcare system at a given time; clinician familiarity with management of patients may vary; and changes in transmissibility and virulence are expected to occur.

The observed variability should inform on the limitations of using observational data during a long-lasting pandemic for management purposes in practice, and also question the use of some variables, such as length of stay in hospital or in ICU, as clinical trial outcomes. This demonstrates the importance of controlling for patient outcome data when designing clinical trials; for example, using our data, assessing a new treatment during the months of March to July will have shown a decrease in CFR from 33%, to 21% that may have been falsely attributed to a treatment effect without a concurrent randomised control.

At the same time, these findings also highlight the need for preparedness and resilience; the crucial importance of pre-positioned observational data collection systems that are adhered upon by a representative number of sites and are maintained for as long as the pandemic lasts; and the need for such capacity to be kept in-between epidemics.

## Data Availability

Data from institutions who have agreed to third party sharing are available to researchers approved by the COIVD-19 Data Platform Data Access Committee. See https://www.iddo.org/covid19/data-sharing/accessing-data for details and the application form.

https://www.iddo.org/covid-19

## Acknowledgments

This work was supported by the UK Foreign, Commonwealth and Development Office and Wellcome [215091/Z/18/Z] and the Bill & Melinda Gates Foundation [OPP1209135]; CIHR Coronavirus Rapid Research Funding Opportunity OV2170359; Grants from Rapid European COVID-19 Emergency Response research (RECOVER) [H2020 project 101003589] and European Clinical Research Alliance on Infectious Diseases (ECRAID) [965313]; The Imperial NIHR Biomedical Research Centre; The Cambridge NIHR Biomedical Research Centre; and Endorsed by the Irish Critical Care-Clinical Trials Group, co-ordinated in Ireland by the Irish Critical Care-Clinical Trials Network at University College Dublin and funded by the Health Research Board of Ireland [CTN-2014-12].

This work uses Data / Material provided by patients and collected by the NHS as part of their care and support #DataSavesLives. The Data / Material used for this research were obtained from ISARIC4C. The COVID-19 Clinical Information Network (CO-CIN) data was collated by ISARIC4C Investigators. Data and Material provision was supported by grants from: the National Institute for Health Research (NIHR; award CO-CIN-01), the Medical Research Council (MRC; grant MC_PC_19059), and by the NIHR Health Protection Research Unit (HPRU) in Emerging and Zoonotic Infections at University of Liverpool in partnership with Public Health England (PHE), (award 200907), Wellcome Trust [Turtle, Lance-fellowship 205228/Z/16/Z], NIHR HPRU in Respiratory Infections at Imperial College London with PHE (award 200927), Liverpool Experimental Cancer Medicine Centre (grant C18616/A25153), NIHR Biomedical Research Centre at Imperial College London (award IS-BRC-1215-20013), and NIHR Clinical Research Network providing infrastructure support.

This work was possible due to the dedication and hard work of the Norwegian SARS-CoV-2 study team, and supported by grants from Research Council of Norway grant no 312780, and a philanthropic donation from Vivaldi Invest A/S owned by Jon Stephenson von Tetzchner; The dedication and hard work of the Groote Schuur Hospital Covid ICU Team, and supported by the Groote Schuur nursing and University of Cape Town registrar bodies coordinated by the Division of Critical Care at the University of Cape Town; and supported by the COVID clinical management team, AIIMS, Rishikesh, India.

Matthew Hall and Christophe Fraser were supported by a Li Ka Shing Foundation award to Christophe Fraser.

