## Supplemental information for "Ten months of temporal variation in the clinical journey of hospitalised patients with COVID-19: an observational cohort"

**Text S1:**

Dependent variables used were:

- Month of COVID-19 admission, or, for question 1, month of symptom onset, as a discrete variable. April was used as the reference category; this is because the first month chronologically, March, was an outlier in many ways and comparisons with that month tended to give universally small *p*-values when compared to other months.
- Sex
- Final outcome (death or discharge)
- Age group (at admission), defined as 0-19, 20-39, 40-59, 60-69, 70-79, or 80+. The 40-59 age group was used as the reference category,
- For questions 2 to 5, either the time from symptom onset to hospital admission, or a variable indicating a nosocomial infection. This was binned as follows: nosocomial infection, 0-6 days, 7-13 days, and 14 days or greater. 0-6 days was the reference category.
- Country. In each analysis, countries with less than 200 patients represented were grouped by continent. Note that, due to differences in recruitment procedures between sites in the consortium, estimated regression coefficients for this variable should not be taken as representative of overall differences in clinical care between countries, nor of demographic differences.
- The four most common symptoms recorded at admission were cough, fatigue, fever, and shortness of breath. For question 1, the number of these that were reported at hospital admission was counted, giving a score from 0 to 4. Missing data on these was counted as

absence. It was excluded from questions 2 to 5 because it proved to be highly collinear with time from onset to admission (see Results).

- The presence or absence of three gastrointestinal symptoms - abdominal pain, diarrhoea, and vomiting - was also recorded on admission. The number of these was counted to give a score from 0 to 3, as above, and used in all analysis except 6.
- A wide variety of comorbidities were also recorded (see table S1). As these data were frequently not recorded, such that treating unknown values as missing would severely decrease the size of the dataset used in the regression analyses, we allowed these variables to take separate values of “Present”, “Absent” and “Unknown”. These were not used in analyses 1 or 6.

**Figure S1:** Proportion of patients entering ICU (a) and time from COVID-19 admission to ICU admission (b) over time. Each line is the proportion (a) or mean value (b) amongst all patients (black, dotted) or patients in each age group (coloured).

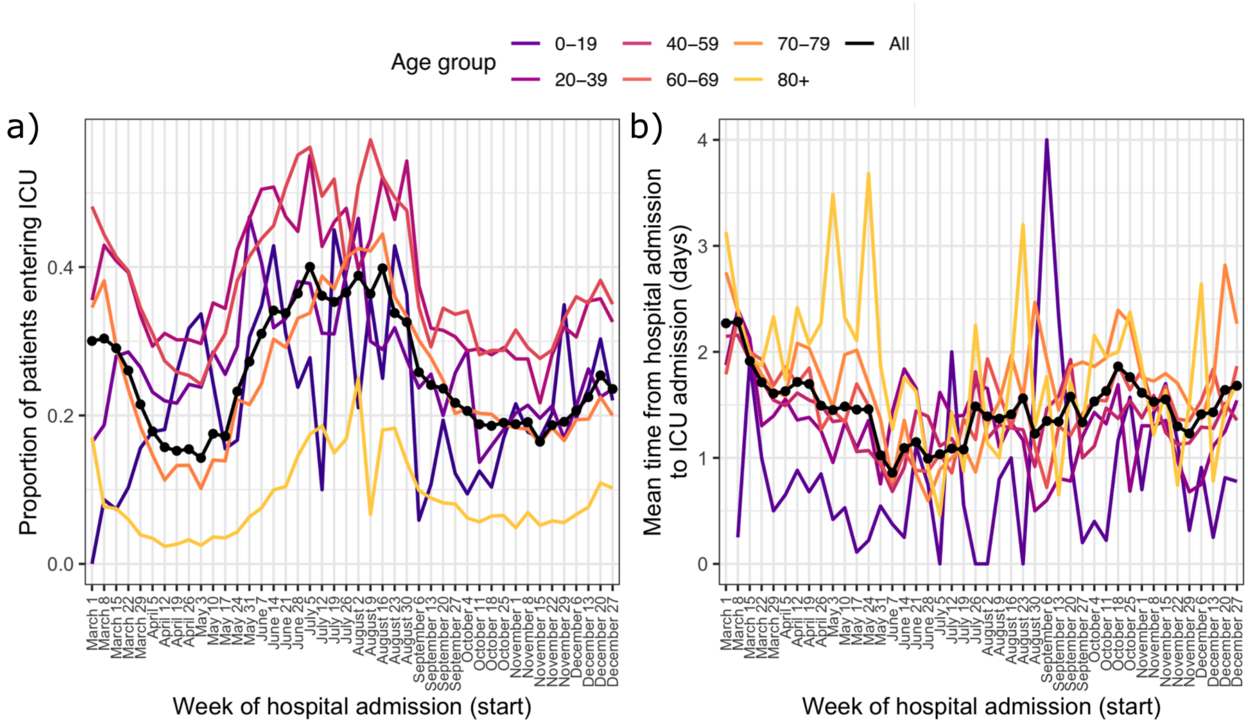

**Figure S2:** Extended version of figure 2, displaying Sankey plots for the patient journey for patients admitted in every month from March to December, subdivided by age group. Bars are presented for the day of admission (A), three and seven days later (A+3 and A+7), and the day after final outcome (O+1).

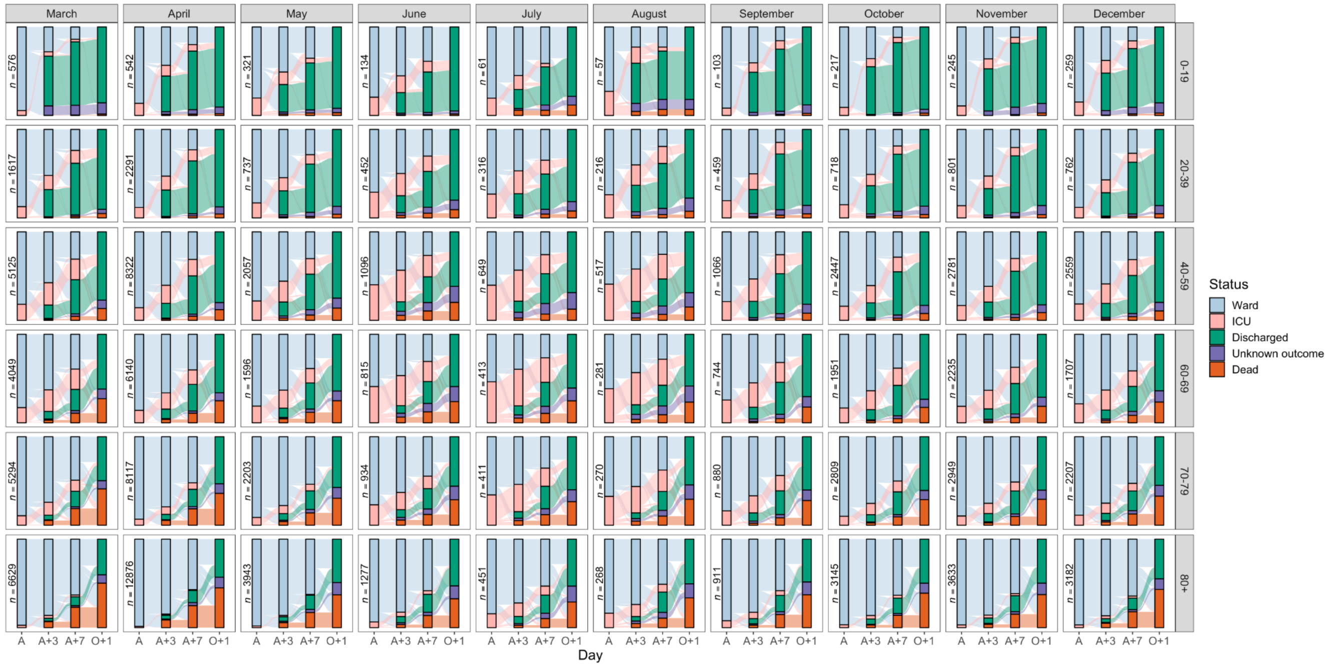

**Figure S3:** Temporal variation in case fatality rate amongst all patients.

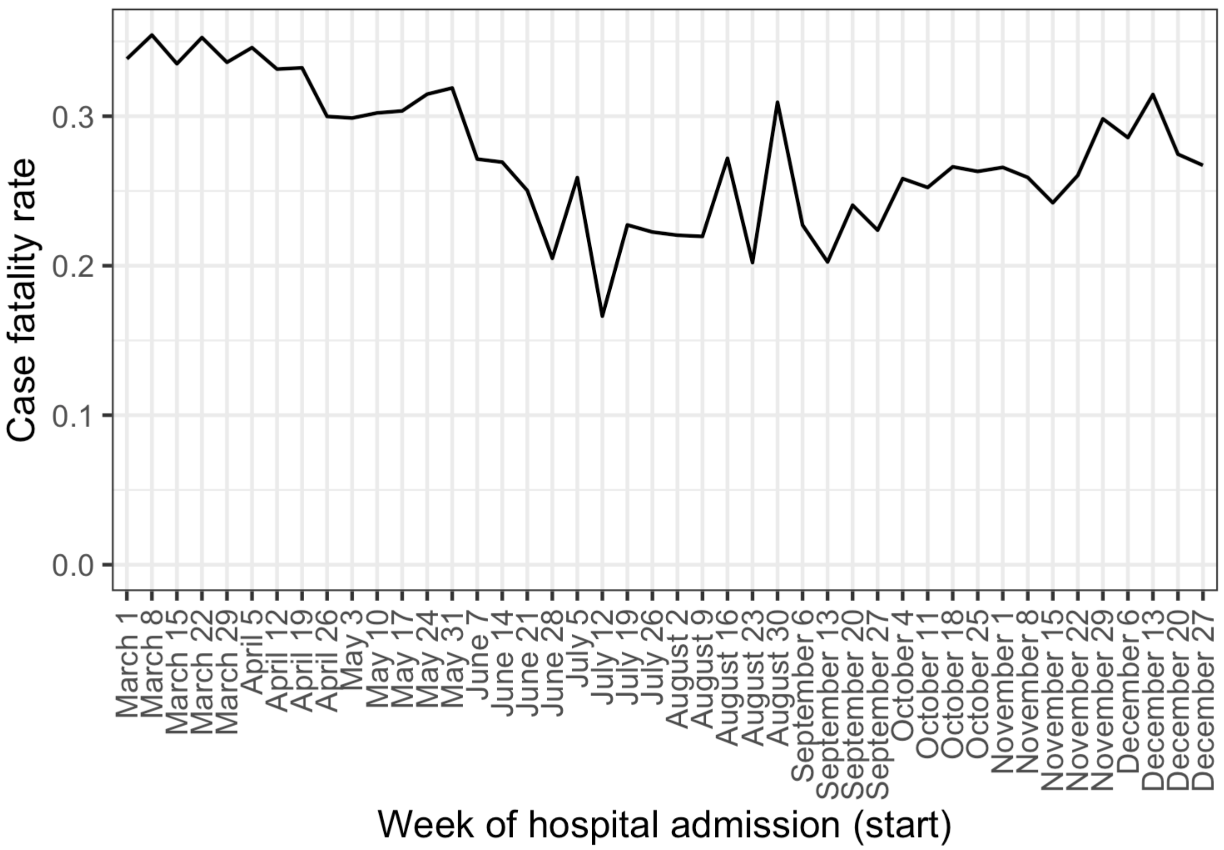

**Figure S4:** Temporal variation in mean time from COVID-19 admission to outcome (death or discharge) amongst all patients.

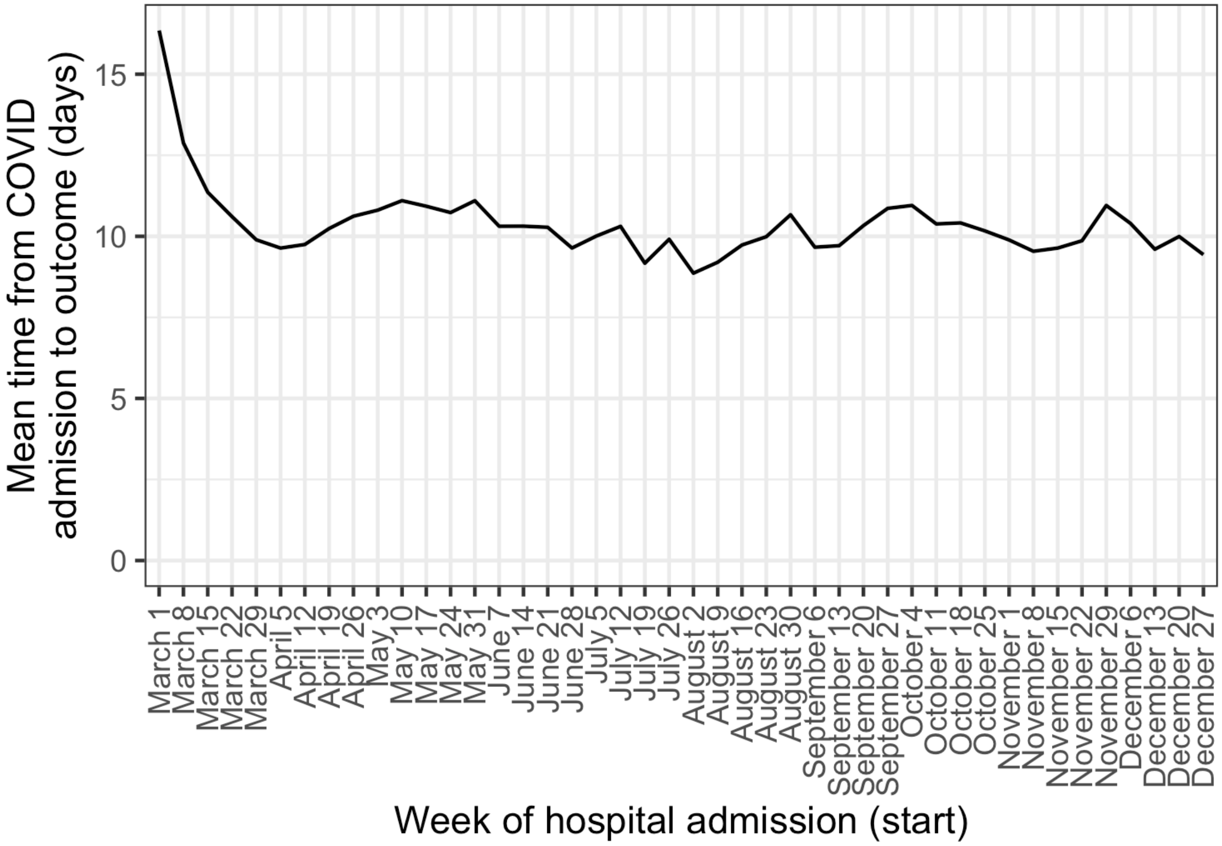

**Figure S5:** Temporal trends in case fatality rate, faceted by ICU/HDU admission and further separated by age group

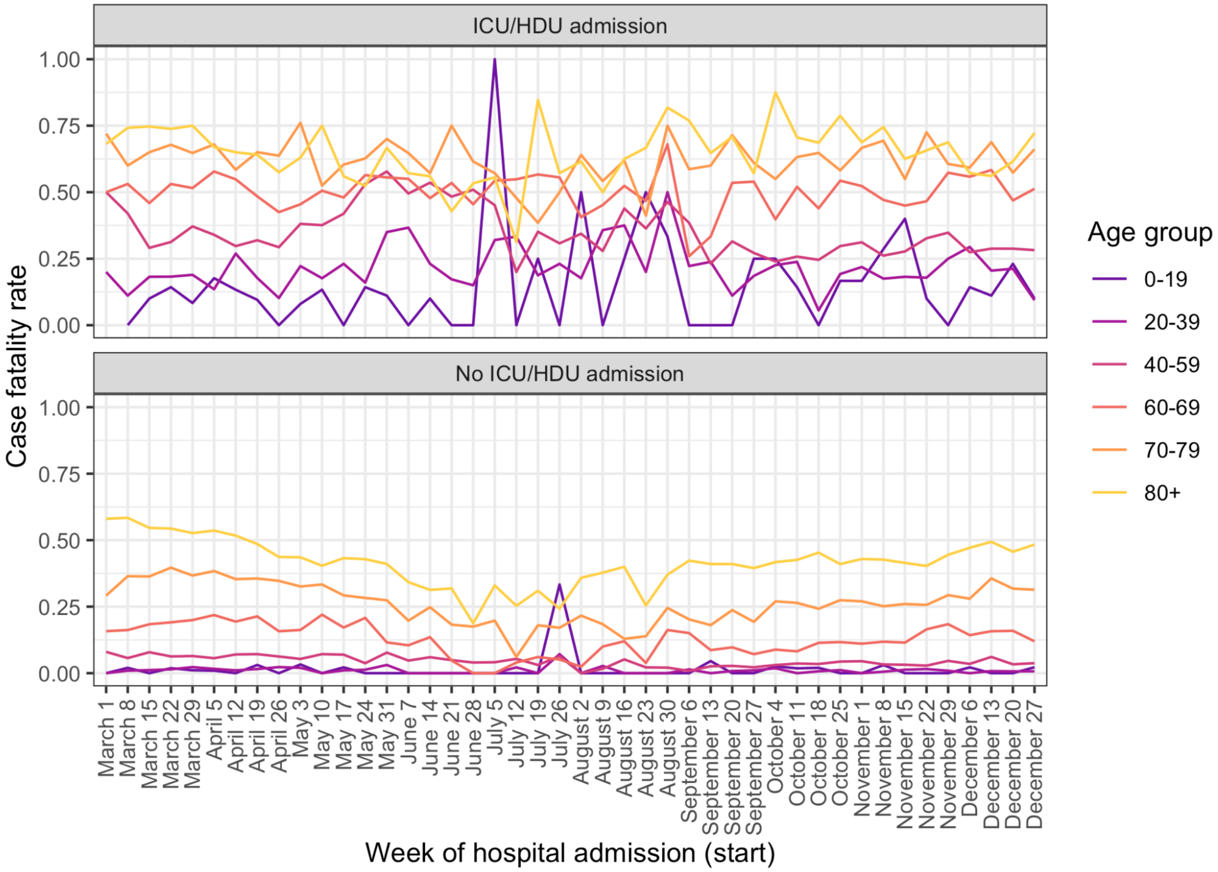

**Figure S6:** Temporal trends in mean time from COVID-19 admission to outcome, faceted by ICU admission and outcome and further separated by age group.

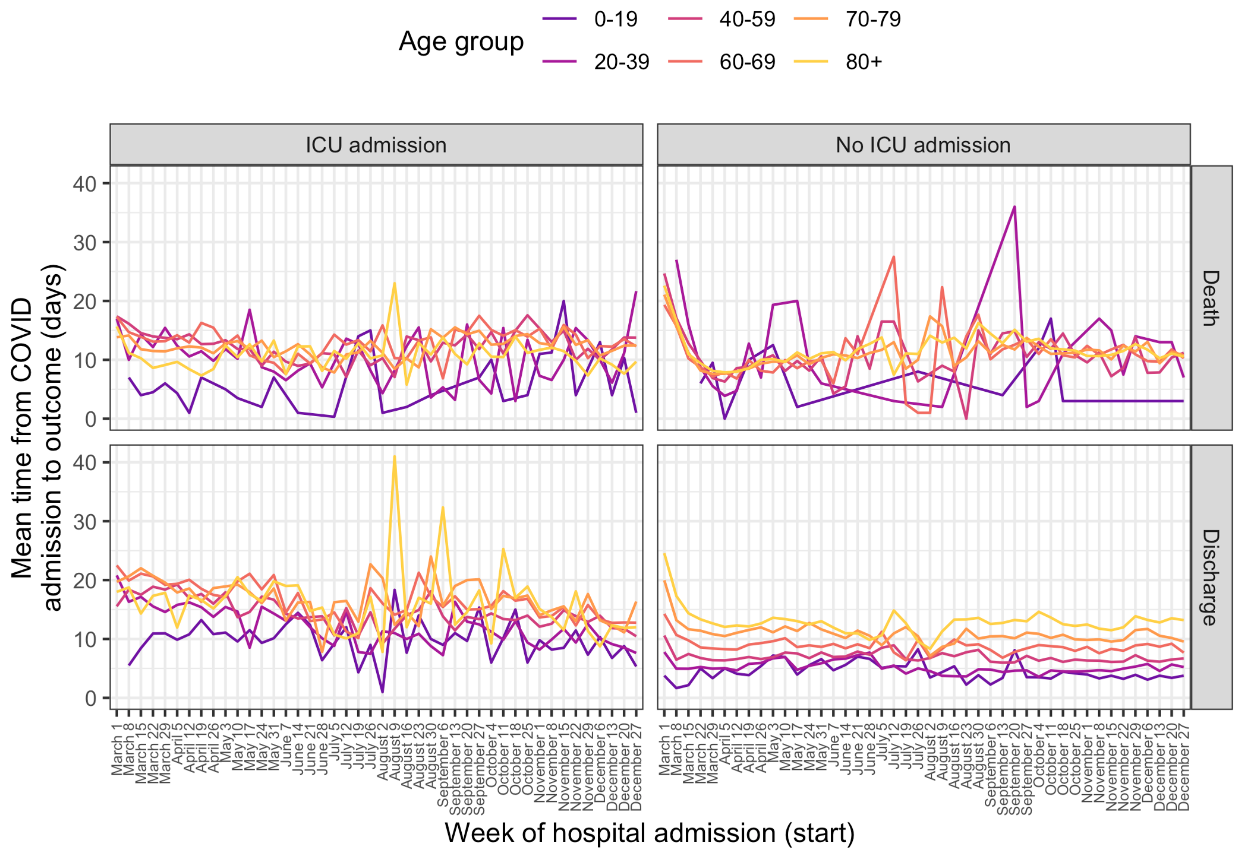

**Table S1:** Extended table of patient characteristics, by month of COVID-19 admission and overall. Raw numbers are given with percentages in brackets.

**Table S2:** Summary of the components of the inpatient journey and their variation over the course of 2020. All time periods are in days. Patients are categorised by month of symptom onset for onset to admission, and by month of COVID admission in all other cases. Patients with COVID admission in 2021, who are included in the analysis of time from onset to admission if their onset date was in 2020, are not listed here as they are excluded from any analysis where the outcome variable is not time from onset to admission. "Outcome" is either death or discharge, and the "admission to outcome" column gives the total length of hospital stay. For all durations, the top 2.5% of values are excluded as potentially misentered.

| Month | Onset to hospital admission | | Proportion entering ICU | COVID-19 admission to ICU | | COVID-19 admission to death | | COVID-19 admission to discharge | | COVID-19 admission to outcome | |
|---|---|---|---|---|---|---|---|---|---|---|---|
| | Mean | SD | | Mean | SD | Mean | SD | Mean | SD | Mean | SD |
| March | 6.82 | 5.15 | 0.262 | 1.79 | 2.35 | 10.6 | 8.41 | 10.9 | 9.71 | 10.8 | 9.29 |
| April | 4.27 | 4.53 | 0.174 | 1.64 | 2.36 | 9.11 | 7.96 | 10.3 | 9.14 | 9.91 | 8.78 |
| May | 4.09 | 4.58 | 0.174 | 1.46 | 2.46 | 10.5 | 8.41 | 11 | 9.62 | 10.9 | 9.28 |
| June | 4.4 | 4.51 | 0.316 | 1.04 | 2.12 | 10.5 | 8.65 | 10.5 | 8.83 | 10.5 | 8.78 |
| July | 4.77 | 4.22 | 0.372 | 1.1 | 2.3 | 11.2 | 8.68 | 9.53 | 8.37 | 9.88 | 8.46 |
| August | 5.49 | 4.6 | 0.377 | 1.4 | 2.49 | 11.6 | 8.82 | 8.88 | 7.88 | 9.44 | 8.16 |
| September | 6.3 | 5.01 | 0.247 | 1.38 | 2.21 | 13.5 | 9.81 | 9.25 | 8.64 | 10.2 | 9.08 |
| October | 5.72 | 4.89 | 0.192 | 1.69 | 2.59 | 12.5 | 8.69 | 9.78 | 8.75 | 10.5 | 8.81 |
| November | 5.17 | 4.75 | 0.183 | 1.49 | 2.5 | 11.7 | 8.38 | 9.1 | 8.08 | 9.78 | 8.24 |
| December | 5.61 | 4.76 | 0.225 | 1.51 | 2.48 | 11.3 | 8 | 9.52 | 8.16 | 10 | 8.15 |

**Table S3:** Prevalence of symptoms amongst patients not presenting with any of cough, shortness of breath, fatigue or fever, by age group. Numbers are percentages (total/total number of patients for whom presence or absence of symptom was reported).

| Symptom | Age group | | | | | |
|---|---|---|---|---|---|---|
| | 0-19 | 20-39 | 40-59 | 60-69 | 70-79 | 80+ |
| Abdominal pain | 40.3 (60/149) | 44.2 (170/385) | 36.4 (223/613) | 30.8 (148/480) | 24.1 (205/849) | 18.2 (306/1682) |
| Ageusia | 8.93 (10/112) | 14.6 (45/308) | 5.5 (26/473) | 2.01 (7/348) | 3.73 (22/590) | 2.15 (25/1161) |
| Anosmia | 7.89 (9/114) | 15.7 (48/305) | 5.5 (26/473) | 2.01 (7/349) | 1.35 (8/592) | 1.11 (13/1167) |
| Bleeding | 1.92 (3/156) | 7.1 (26/366) | 5.18 (31/599) | 4.09 (19/464) | 4.86 (40/823) | 6.25 (103/1648) |
| Confusion | 9.8 (15/153) | 7.65 (29/379) | 25.1 (159/634) | 42 (209/498) | 59.4 (551/928) | 73.1 (1381/1889) |
| Conjunctivitis | 0 (0/151) | 0.57 (2/351) | 0.344 (2/581) | 0.671 (3/447) | 0.252 (2/793) | 0.255 (4/1570) |
| Diarrhoea | 17.4 (28/161) | 16.6 (62/373) | 20.5 (125/609) | 25.7 (122/475) | 20.5 (174/850) | 16.2 (272/1684) |
| Ear pain | 1.79 (2/112) | 3.2 (9/281) | 0.98 (5/510) | 0 (0/404) | 0.409 (3/734) | 0.412 (6/1458) |
| Headache | 13.1 (18/137) | 26.4 (96/364) | 21.9 (131/598) | 14.3 (64/447) | 7.4 (59/797) | 4.61 (72/1561) |
| Lymphadenopathy | 1.33 (2/150) | 0.283 (1/353) | 0.515 (3/582) | 0.891 (4/449) | 0.879 (7/796) | 0.506 (8/1580) |
| Myalgia | 5.07 (7/138) | 12 (43/359) | 11.5 (67/582) | 9.26 (41/443) | 6.42 (50/779) | 3.69 (56/1517) |
| Rash | 9.55 (15/157) | 1.98 (7/354) | 1.37 (8/586) | 2.2 (10/454) | 0.753 (6/797) | 1.58 (25/1587) |
| Runny nose | 14.7 (22/150) | 6.46 (23/356) | 3.99 (23/577) | 0.456 (2/439) | 0.641 (5/780) | 0.846 (13/1537) |
| Seizures | 12.3 (19/154) | 3.72 (14/376) | 7.51 (45/599) | 7.48 (35/468) | 5.53 (46/832) | 4.11 (68/1655) |
| Ulcers | 1.6 (2/125) | 0.697 (2/287) | 1.86 (10/538) | 1.9 (8/422) | 4.74 (37/781) | 4.08 (64/1567) |
| Vomiting | 50 (82/164) | 42.9 (166/387) | 47.1 (296/628) | 40.9 (198/484) | 30.5 (265/870) | 21.5 (372/1728) |
| Wheezing | 1.96 (3/153) | 1.12 (4/358) | 1.86 (11/592) | 2.37 (11/464) | 2.08 (17/817) | 2.67 (43/1611) |

**Table S4:** Results of a linear regression analysis identifying variables associated with time from symptom onset to hospital admission. The dependent variable was log-transformed after the addition of 1 day to prevent log-transforming zero values. Coefficients have been transformed to reflect the percentage increase or decrease in the number of days to admission, compared to the reference category for categorical variables, or per one point increase for discrete or continuous variables. Countries with less than 200 patients in the dataset have been grouped by region.

| | % increase in time to admission (days) |
|---|---|
| Month of symptom onset (ref: April) | |
| March | 44.548*** (42.455, 46.673) |
| May | −3.550** (−5.651, −1.404) |
| June | −4.714** (−7.700, −1.631) |
| July | 0.522 (−3.800, 5.038) |
| August | 21.353*** (15.762, 27.214) |
| September | 43.579*** (39.760, 47.501) |
| October | 31.937*** (29.404, 34.520) |
| November | 21.176*** (18.777, 23.623) |
| December | 29.025*** (26.496, 31.604) |
| Number of common symptoms (max 4) | 25.015*** (24.406, 25.626) |
| Number of GI symptoms (max 3) | 9.753*** (8.981, 10.530) |
| Age group (ref: 40-59) | |
| 0-19 | −33.357*** (−35.905, −30.708) |
| 20-39 | −9.598*** (−11.562, −7.591) |
| 60-69 | −4.183*** (−5.736, −2.604) |
| 70-79 | −17.013*** (−18.317, −15.688) |
| 80+ | −29.671*** (−30.768, −28.556) |
| Sex (ref: Female) | |
| Male | 4.360*** (3.262, 5.470) |
| Final outcome (ref: death) | |
| Discharge | 13.035*** (11.638, 14.450) |
| Country (ref: Africa and Middle East - grouped) | |
| Asia - grouped | 4.410 (−13.404, 25.890) |
| Belgium | −0.738 (−10.530, 10.126) |
| Brazil | 41.089*** (26.179, 57.761) |
| Canada | 4.538 (−4.555, 14.497) |
| Colombia | 28.348*** (13.811, 44.741) |
| Europe - grouped | 11.503 (−2.499, 27.515) |
| France | −4.792 (−13.088, 4.296) |
| India | −2.474 (−13.640, 10.136) |
| Indonesia | −2.602 (−13.720, 9.947) |
| Ireland | −1.823 (−11.805, 9.289) |
| Israel | 14.266* (1.045, 29.216) |
| Italy | 15.956* (3.105, 30.409) |
| Nepal | 36.831*** (15.354, 62.307) |
| Netherlands | 8.396 (−1.456, 19.233) |
| Norway | 14.960* (1.320, 30.436) |
| Pakistan | 57.796*** (43.445, 73.584) |
| Peru | 84.213*** (60.299, 111.695) |
| Poland | −0.486 (−14.231, 15.462) |
| Portugal | 11.790* (0.121, 24.819) |
| Romania | 24.092*** (11.899, 37.613) |
| South and Central America and Caribbean - grouped | −1.319 (−15.320, 14.995) |
| Spain | −8.218 (−19.285, 4.366) |
| United Kingdom | −10.399* (−17.631, −2.533) |
| United States of America | −7.490 (−15.999, 1.882) |
| (Intercept) | 138.760*** (119.063, 160.228) |
| Observations | 99,729 |
| *Note:* | *p<0.05; **p<0.01; ***p<0.001 |

**Table S5:** Results of logistic regression identifying variables associated with ICU admission. Countries with less than 200 patients in the dataset have been grouped by region.

| | Odds ratio (95% CI) |
|---|---|
| Month of COVID admission (ref: April) | |
| March | 1.346*** (1.278, 1.418) |
| May | 1.006 (0.927, 1.093) |
| June | 1.317*** (1.169, 1.482) |
| July | 1.808*** (1.550, 2.108) |
| August | 1.808*** (1.530, 2.136) |
| September | 1.413*** (1.277, 1.563) |
| October | 1.243*** (1.157, 1.335) |
| November | 1.101** (1.025, 1.182) |
| December | 1.342*** (1.248, 1.444) |
| Number of GI symptoms (max 3) | 0.935*** (0.912, 0.959) |
| Age group (ref: 40-59) | |
| 0-19 | 0.869* (0.761, 0.994) |
| 20-39 | 0.755*** (0.700, 0.815) |
| 60-69 | 0.966 (0.917, 1.017) |
| 70-79 | 0.506*** (0.477, 0.536) |
| 80+ | 0.124*** (0.115, 0.134) |
| Sex (ref: Female) | |
| Male | 1.527*** (1.466, 1.589) |
| Days from symptom onset to hospital admission (ref: 0-6): | |
| Nosocomial infection | 0.679*** (0.622, 0.741) |
| 7-13 | 1.363*** (1.307, 1.422) |
| 14+ | 1.280*** (1.200, 1.365) |
| Final outcome (ref: death) | |
| Discharge | 0.186*** (0.177, 0.195) |
| Pregnant (ref: no) | |
| Yes | 0.917 (0.733, 1.146) |
| Unknown | 0.794** (0.678, 0.931) |
| Comorbidities (ref: absent) | |
| Asthma | |
| Present | 1.044 (0.989, 1.102) |
| Unknown | 0.997 (0.833, 1.193) |
| Chronic cardiac disease | |
| Present | 0.698*** (0.663, 0.735) |
| Unknown | 0.789** (0.679, 0.917) |
| Chronic haemotologic disease | |
| Present | 0.954 (0.857, 1.061) |
| Unknown | 1.997*** (1.647, 2.420) |
| Chronic kidney disease | |
| Present | 0.675*** (0.632, 0.721) |
| Unknown | 0.662*** (0.546, 0.804) |
| Chronic neurological disorder | |
| Present | 0.570*** (0.528, 0.614) |
| Unknown | 0.778* (0.636, 0.952) |
| Chronic pulmonary disease | |
| Present | 0.632*** (0.595, 0.672) |
| Unknown | 1.029 (0.855, 1.237) |
| Dementia | |
| Present | 0.210*** (0.185, 0.238) |
| Unknown | 0.591*** (0.504, 0.693) |
| Diabetes | |
| Present | 1.054 (0.999, 1.112) |
| Unknown | 0.833*** (0.769, 0.902) |
| HIV/AIDS | |
| Present | 1.459** (1.135, 1.875) |
| Unknown | 0.977 (0.892, 1.071) |
| Hypertension | |
| Present | 1.262*** (1.204, 1.324) |
| Unknown | 1.237*** (1.170, 1.308) |
| Liver disease | |
| Present | 0.747*** (0.671, 0.833) |
| Unknown | 2.648*** (2.238, 3.133) |
| Malignant neoplasm | |
| Present | 0.613*** (0.567, 0.662) |
| Unknown | 0.548*** (0.449, 0.669) |
| Malnutrition | |
| Present | 0.889 (0.759, 1.041) |
| Unknown | 0.794*** (0.719, 0.876) |
| Obesity | |
| Present | 1.739*** (1.653, 1.830) |
| Unknown | 0.883*** (0.823, 0.948) |
| Rheumatological disorder | |
| Present | 1.052 (0.981, 1.128) |
| Unknown | 1.598*** (1.361, 1.877) |
| Smoking | |
| Present | 0.819*** (0.749, 0.896) |
| Unknown | 0.995 (0.955, 1.036) |
| Country (ref: Asia and Oceania - grouped) | |
| Africa and Middle East - grouped | $9.11\times10^{6}$ ($1.59\times10^{-100}$, $5.23\times10^{113}$) |
| Belgium | 0.944 (0.534, 1.669) |
| Brazil | 1.300 (0.723, 2.339) |
| Canada | 1.261 (0.722, 2.205) |
| Colombia | 4.905*** (2.658, 9.051) |
| Europe - grouped | 0.893 (0.475, 1.680) |
| France | 0.608 (0.348, 1.060) |
| India | 1.283 (0.709, 2.321) |
| Indonesia | 4.415** (1.815, 10.741) |
| Ireland | 0.601 (0.339, 1.065) |
| Israel | 0.278*** (0.146, 0.528) |
| Italy | 0.700 (0.386, 1.269) |
| Nepal | $1.46\times10^{7}$ ($2.37\times10^{-161}$, $8.97\times10^{174}$) |
| Netherlands | 0.598 (0.341, 1.047) |
| Norway | 0.406** (0.220, 0.750) |
| Pakistan | $1.13\times10^{7}$ ($9.49\times10^{-47}$, $1.35\times10^{60}$) |
| Peru | 0.080*** (0.037, 0.174) |
| Poland | 0.028*** (0.009, 0.089) |
| Portugal | 18.849*** (10.047, 35.365) |
| Romania | 0.057*** (0.029, 0.111) |
| South and Central America and Caribbean - grouped | 3.116** (1.464, 6.630) |
| Spain | 0.946 (0.515, 1.739) |
| United Kingdom | 0.319*** (0.185, 0.550) |
| United States of America | 6.712*** (3.779, 11.919) |
| (Intercept) | 2.170** (1.252, 3.761) |
| Observations | 105,765 |

*Note:* *p<0.05; **p<0.01; ***p<0.001

**Table S6:** Results of a linear regression analysis identifying variables associated with time from to hospital admission to ICU admission, amongst patients with any ICU admission. The dependent variable was log-transformed after the addition of 1 day to prevent log-transforming zero values. Coefficients have been transformed to reflect the percentage increase or decrease in the number of days to admission, compared to the reference category for categorical variables, or per one point increase for discrete or continuous variables. Countries with less than 200 patients in the dataset have been grouped by region.

| | % increase in time to ICU admission (95% CI) |
|---|---|
| Month of COVID admission (ref: April) | |
| March | 7.076*** (4.072, 10.167) |
| May | 0.151 (−4.007, 4.488) |
| June | 0.823 (−4.105, 6.005) |
| July | −1.809 (−7.622, 4.371) |
| August | 2.701 (−4.103, 9.988) |
| September | 1.542 (−3.727, 7.098) |
| October | 6.989*** (2.783, 11.367) |
| November | 4.033 (−0.085, 8.321) |
| December | 8.277*** (3.766, 12.985) |
| Number of GI symptoms (max 3) | 8.978*** (7.488, 10.488) |
| Age group (ref: 40-59) | |
| 0-19 | −31.991*** (−37.069, −26.503) |
| 20-39 | −4.517* (−8.268, −0.612) |
| 60-69 | 2.257 (−0.364, 4.948) |
| 70-79 | 4.197** (1.161, 7.324) |
| 80+ | 3.740 (−0.621, 8.293) |
| Sex (ref: Female) | |
| Male | −0.062 (−2.232, 2.156) |
| Days from symptom onset to hospital admission (ref: 0-6): | |
| Nosocomial infection | 65.993*** (54.431, 78.419) |
| 7-13 | −9.197*** (−11.138, −7.214) |
| 14+ | −14.872*** (−17.760, −11.883) |
| Final outcome (ref: death) | |
| Discharge | −1.940 (−4.038, 0.204) |
| Pregnant (ref: no) | |
| Yes | 17.541* (3.417, 33.594) |
| Unknown | 4.610 (−4.316, 14.368) |
| Comorbidities (ref: absent) | |
| Asthma | |
| Present | 6.962*** (3.787, 10.235) |
| Unknown | 8.473 (−3.331, 21.717) |
| Chronic cardiac disease | |
| Present | 1.885 (−1.022, 4.876) |
| Unknown | −3.563 (−11.393, 4.958) |
| Chronic haemotologic disease | |
| Present | 20.567*** (12.761, 28.914) |
| Unknown | 0.578 (−10.274, 12.742) |
| Chronic kidney disease | |
| Present | 9.842*** (5.896, 13.936) |
| Unknown | 7.458 (−5.296, 21.930) |
| Chronic neurological disorder | |
| Present | −0.705 (−5.042, 3.830) |
| Unknown | −9.739 (−20.980, 3.101) |
| Chronic pulmonary disease | |
| Present | 1.148 (−2.218, 4.629) |
| Unknown | −1.448 (−12.176, 10.591) |
| Dementia | |
| Present | −7.548 (−14.999, 0.555) |
| Unknown | −7.872 (−16.642, 1.822) |
| Diabetes | |
| Present | −3.457* (−6.111, −0.729) |
| Unknown | −0.407 (−4.899, 4.298) |
| HIV/AIDS | |
| Present | 6.189 (−6.934, 21.163) |
| Unknown | 9.536*** (4.222, 15.120) |
| Hypertension | |
| Present | −1.370 (−3.753, 1.073) |
| Unknown | −4.190** (−7.088, −1.203) |
| Liver disease | |
| Present | 2.895 (−3.553, 9.773) |
| Unknown | −11.076* (−19.342, −1.962) |
| Malignant neoplasm | |
| Present | 3.292 (−1.392, 8.198) |
| Unknown | 0.355 (−11.095, 13.280) |
| Malnutrition | |
| Present | 1.622 (−7.354, 11.467) |
| Unknown | 2.908 (−3.201, 9.402) |
| Obesity | |
| Present | −7.547*** (−9.920, −5.111) |
| Unknown | −0.827 (−4.959, 3.485) |
| Rheumatological disorder | |
| Present | 4.158 (−0.302, 8.818) |
| Unknown | 5.937 (−4.679, 17.735) |
| Smoking | |
| Present | −4.914* (−9.474, −0.125) |
| Unknown | −6.309*** (−8.378, −4.194) |
| Country (ref: Africa and Middle East - grouped) | |
| Asia and Oceania - grouped | −10.453 (−25.109, 7.071) |
| Belgium | −5.612 (−16.088, 6.172) |
| Brazil | −17.749** (−28.321, −5.619) |
| Canada | −17.040** (−25.873, −7.153) |
| Colombia | −25.190*** (−33.339, −16.044) |
| Europe - grouped | −1.511 (−13.528, 12.175) |
| France | 21.404*** (8.256, 36.148) |
| India | −32.270*** (−40.683, −22.664) |
| Indonesia | −9.011 (−18.856, 2.028) |
| Ireland | −2.785 (−15.195, 11.441) |
| Italy | −3.152 (−14.690, 9.947) |
| Nepal | −51.762*** (−58.636, −43.746) |
| Netherlands | −9.352 (−19.535, 2.120) |
| Pakistan | −50.529*** (−55.163, −45.416) |
| Portugal | −5.375 (−15.030, 5.376) |
| South and Central America and Caribbean - grouped | −0.837 (−13.276, 13.386) |
| Spain | −7.482 (−19.530, 6.369) |
| United Kingdom | −1.617 (−10.176, 7.757) |
| United States of America | −17.895*** (−25.293, −9.764) |
| (Intercept) | 118.055*** (97.726, 140.474) |
| Observations | 18,686 |
| *Note:* | *p<0.05; **p<0.01; ***p<0.001 |

**Table S7:** Expanded version of table 3, including hazard ratios for the "unknown" class for comorbidities. These are largely nuisance parameters, and are included for completeness only.

| Variable | Hazard ratio (Death) | Hazard ratio (Discharge) |
|---|---|---|
| Month of COVID admission (ref: May) | | |
| March | 1.27*** (1.21, 1.34) | 0.903*** (0.872, 0.935) |
| April | 1.32*** (1.26, 1.39) | 1 (0.973, 1.04) |
| June | 0.822*** (0.746, 0.905) | 1.11*** (1.06, 1.17) |
| July | 0.582*** (0.486, 0.697) | 1.27*** (1.19, 1.36) |
| August | 0.598*** (0.483, 0.74) | 1.2*** (1.11, 1.31) |
| September | 0.778*** (0.701, 0.864) | 1.07* (1.01, 1.12) |
| October | 0.888*** (0.829, 0.951) | 1.12*** (1.08, 1.16) |
| November | 0.997 (0.927, 1.07) | 1.27*** (1.22, 1.32) |
| Age group (ref: 40-59) | | |
| 0-19 | 0.427*** (0.261, 0.697) | 1.69*** (1.6, 1.8) |
| 20-39 | 0.355*** (0.272, 0.463) | 1.4*** (1.35, 1.45) |
| 60-69 | 2.09*** (1.92, 2.28) | 0.692*** (0.673, 0.711) |
| 70-79 | 3.23*** (2.99, 3.49) | 0.49*** (0.477, 0.504) |
| 80+ | 4.37*** (4.05, 4.71) | 0.352*** (0.343, 0.362) |
| Past or current ICU admission | 2.78*** (2.45, 3.15) | 0.28*** (0.256, 0.306) |
| Sex (ref: Female) | | |
| Male | 1.21*** (1.18, 1.24) | 0.916*** (0.901, 0.932) |
| Number of GI symptoms (max 3) | 0.882*** (0.865, 0.899) | 1.06*** (1.04, 1.07) |
| Days from symptom onset to hospital admission (ref: 0-6): | | |
| Nosocomial infection | 0.74*** (0.71, 0.77) | 0.466*** (0.451, 0.483) |
| 7-13 | 0.868*** (0.84, 0.896) | 1.21*** (1.19, 1.23) |
| 14+ | 0.888*** (0.848, 0.93) | 1.19*** (1.15, 1.22) |
| Pregnant (ref: no) | | |
| Yes | 0.788 (0.486, 1.28) | 1.5*** (1.38, 1.64) |
| Unknown | 0.949 (0.748, 1.2) | 1.11** (1.04, 1.19) |
| Comorbidities (ref: absent) | | |
| Asthma | | |
| Present | 0.954* (0.917, 0.993) | 1.04** (1.01, 1.06) |
| Unknown | 1.1 (0.991, 1.21) | 1.04 (0.966, 1.13) |
| Chronic cardiac disease | | |
| Present | 1.06 (0.872, 1.29) | 0.876 (0.76, 1.01) |
| Unknown | 0.903 (0.694, 1.17) | 0.748** (0.627, 0.893) |
| Chronic haemotologic disease | | |
| Present | 1.1** (1.03, 1.16) | 0.835*** (0.798, 0.875) |
| Unknown | 0.919 (0.815, 1.04) | 0.781*** (0.709, 0.86) |
| Chronic kidney disease | | |
| Present | 1.25*** (1.21, 1.29) | 0.839*** (0.817, 0.862) |
| Unknown | 0.986 (0.893, 1.09) | 1.16*** (1.07, 1.26) |
| Chronic neurological disorder | | |
| Present | 1.11*** (1.07, 1.15) | 0.731*** (0.71, 0.753) |
| Unknown | 0.931 (0.841, 1.03) | 1.1* (1.01, 1.19) |
| Chronic pulmonary disease | | |
| Present | 1.19*** (1.15, 1.23) | 0.874*** (0.852, 0.896) |
| Unknown | 0.997 (0.903, 1.1) | 1.02 (0.939, 1.1) |
| Dementia | | |
| Present | 1.14*** (1.1, 1.18) | 0.784*** (0.759, 0.81) |
| Unknown | 0.95 (0.879, 1.03) | 1.01 (0.945, 1.07) |
| Diabetes | | |
| Present | 1.1*** (1.07, 1.14) | 0.941*** (0.918, 0.964) |
| Unknown | 1.09*** (1.04, 1.15) | 0.944** (0.911, 0.978) |
| HIV/AIDS | | |
| Present | 1.16 (0.941, 1.44) | 0.884 (0.777, 1.01) |
| Unknown | 0.971 (0.909, 1.04) | 1.05* (1.01, 1.1) |
| Hypertension | | |
| Present | 0.978 (0.949, 1.01) | 0.981 (0.961, 1) |
| Unknown | 1.1*** (1.06, 1.13) | 1.03* (1, 1.05) |
| Liver disease | | |
| Present | 1.19*** (1.11, 1.27) | 0.773*** (0.737, 0.812) |
| Unknown | 1.04 (0.935, 1.16) | 0.748*** (0.687, 0.815) |
| Malignant neoplasm | | |
| Present | 1.18*** (1.14, 1.23) | 0.818*** (0.792, 0.845) |
| Unknown | 0.992 (0.897, 1.1) | 1.19*** (1.1, 1.29) |
| Malnutrition | | |
| Present | 1.07 (0.996, 1.15) | 0.768*** (0.721, 0.819) |
| Unknown | 1.04 (0.991, 1.1) | 0.967 (0.93, 1) |
| Obesity | | |
| Present | 1.08*** (1.04, 1.13) | 0.896*** (0.873, 0.919) |
| Unknown | 1.08*** (1.04, 1.13) | 1.08*** (1.05, 1.11) |
| Rheumatological disorder | | |
| Present | 0.948** (0.911, 0.986) | 0.977 (0.949, 1.01) |
| Unknown | 1.01 (0.924, 1.1) | 0.909** (0.847, 0.977) |
| Smoking | | |
| Present | 1.03 (0.965, 1.09) | 0.854*** (0.822, 0.887) |
| Unknown | 1.08*** (1.05, 1.11) | 0.907*** (0.89, 0.923) |
| Interaction: Past or current ICU admission × month of admission (ref: May) | | |
| March | 0.861** (0.773, 0.959) | 0.836*** (0.761, 0.918) |
| April | 0.79*** (0.711, 0.878) | 0.82*** (0.748, 0.898) |
| June | 1.02 (0.87, 1.19) | 0.992 (0.87, 1.13) |
| July | 1.32* (1.04, 1.67) | 1.14 (0.973, 1.34) |
| August | 1.34* (1.02, 1.75) | 1.13 (0.943, 1.35) |
| September | 0.971 (0.814, 1.16) | 0.979 (0.854, 1.12) |
| October | 1.04 (0.904, 1.19) | 0.97 (0.865, 1.09) |
| November | 0.953 (0.826, 1.1) | 0.895 (0.79, 1.01) |
| Interaction: Past or current ICU admission × age group (ref: 40-59) | | |
| 0-19 | 1.2 (0.655, 2.21) | 1.45*** (1.25, 1.68) |
| 20-39 | 1.96*** (1.46, 2.64) | 1.05 (0.965, 1.13) |
| 60-69 | 0.72*** (0.647, 0.8) | 1.06* (1, 1.13) |
| 70-79 | 0.697*** (0.631, 0.77) | 1.31*** (1.21, 1.41) |
| 80+ | 0.742*** (0.663, 0.829) | 2.37*** (2.11, 2.65) |

*Note:* *p<0.05; **p<0.01; ***p<0.001

**CRediT author statement (based on Brand et al., 2015, doi: 10.1087/20150211):**

**Conceptualization (this analysis):** Carson, Gail; Hall, Matthew; Olliaro, Piero L.; Rojek, Amanda.
**Conceptualization (study):** Baillie, J. Kenneth; Carson, Gail; Dunning, Jake; Horby, Peter; Merson, Laura; Semple, Malcolm G. Methodology: Hall, Matthew; Donnelly, Christl A.; Kartsonaki, Christiana.
**Software and formal analysis:** Dankwa, Emmanuelle; Hall, Matthew; Kartsonaki, Christiana; McLean, Kenneth A.; Pritchard, Mark.
**Data curation:** Citarella, Barbara Wanjiru; Kelly, Sadie; Kennon, Kalynn; Lee, James; Merson, Laura; Plotkin, Daniel; Smith, Sue; Strudwick, Samantha.
**Administration:** Citarella, Barbara Wanjiru; Merson, Laura.
**Writing - original draft:** Baruch, Joaquín; Dagens, Andrew; Dunning, Jake; Hall, Matthew; Kartsonaki, Christiana; Olliaro, Piero L.; Rojek, Amanda. Visualization: Hall, Matthew.
**Writing - review and editing:** All authors.
**Data Contributors- including Investigation, Supervision, Resources, Project administration and Funding acquisition:** Abdukahil, Sheryl Ann; Abe, Ryuzo; Abel, Laurent; Absil, Lara; Acharya, Subhash; Acker, Andrew; Adachi, Shingo; Adam, Elisabeth; Adrião, Diana; Ageel, Saleh Al; Aguirre, Emilio; Ain, Quratul; Ainscough, Kate; Ait Hssain, Ali; Ait Tamlihat, Younes; Akimoto, Takako; Akmal, Ernita; Al Qasim, Eman; Al-dabbous, Tala; Al-Fares, Abdulrahman; Alalqam, Razi; Alam, Tanvir; Alegre, Cynthia; Alex, Beatrice; Alexandre, Kévin; Alfoudri, Huda; Ali Shah, Naseem; Alidjnou, Kazali Enagnon; Aliudin, Jeffrey; Allavena, Clotilde; Allou, Nathalie; Altaf, Aneela; Alves, João; Alves, João Melo; Alves, Rita; Amaral, Maria; Ammerlaan, Heidi; Ampaw, Phoebe; Andini, Roberto; Andrejak, Claire; Angheben, Andrea; Angoulvant, François; Ansart, Séverine; Antonelli, Massimo; Antunes de Brito, Carlos Alexandre; Apriyana, Ardiyan; Arabi, Yaseen; Aragao, Irene; Arali, Rajeshwari; Arancibia, Francisco; Arcadipane, Antonio; Archambault, Patrick; Arenz, Lukas; Arlet, Jean-Benoît; Arnold-Day, Christel; Arora, Lovkesh; Arora, Rakesh; Artaud-Macari, Elise; Aryal, Diptesh; Aryal, Diptesh; Asaki, Motohiro; Asensio, Angel; Assie, Jean Baptiste; Atique, Anika; Attanyake, AM Udara Lakshan; Auchabie, Johann; Aumaitre, Hugues; Auvet, Adrien; Azemar, Laurène; Azhari, Taufik; Azoulay, Cecile; Babin, Camille; Bach, Benjamin; Bachelet, Delphine; Badr, Claudine; Baena, Jose; Bailey, Cassandra; Baillie, J. Kenneth; Bak, Erica; Bakakos, Agamemnon; Baker, Andrew; Bal, Andriy; Banheiro, Bruno; Bani-Sadr, Firouzé; Barbalho, Renata; Barclay, Wendy S.; Barnikel, Michaela; Barrasa, Helena; Barrelet, Audrey; Barrigoto, Cleide; Bartoli, Marie; Bartone, Cheryl; Baruch, Joaquín; Basmaci, Romain; Battaglini, Denise; Bauer, Jules; Bautista, Diego; Beane, Abigail; Beane, Abigail; Bedossa, Alexandra; Behilill, Sylvie; Beljantsev, Aleksandr; Bellemare, David; Beltrame, Anna; Beluze, Marine; Benech, Nicolas; Benkerrou, Dehbia; Bennett, Suzanne; Bento, Luís; Berdal, Jan-Erik; Bergeaud, Delphine; Bernal Salvador, Gabriela; Bernal Sobrino, José Luis; Bertolino, Lorenzo; Bessis, Simon; Betz, Adam; Bevilcaqua, Sybille; Bezulier, Karine; Bhatt, Amar; Bhavsar, Krishna; Bianchi, Isabella; Bianco, Claudia; Bikram Singh, Moirangthem; Bin Humaid, Felwa; Bissell, Erin; Bissuel, François; Biston, Patrick; Bitker, Laurent; Blanco-Schweizer, Pablo; Bloos, Frank; Blot, Mathieu; Boccia, Filomena; Bodenes, Laetitia; Boethel, Carl; Bogaarts, Alice; Bogaert, Debby; Boivin, Anne-Hélène; Bolze, Pierre-Adrien; Bompart, François; Borges, Diogo; Borie, Raphaël; Bosse,

Hans Martin; Botelho-Nevers, Elisabeth; Bouadma, Lila; Bouchaud, Olivier; Bouchez, Sabelline; Bouhmani, Dounia; Bouhour, Damien; Bouiller, Kévin; Bouillet, Laurence; Bouisse, Camile; Boureau, Anne-Sophie; Bouscambert, Maude; Bousquet, Aurore; Bouziotis, Jason; Boxma, Bianca; Boyer-Besseyre, Marielle; Boylan, Maria; Brack, Matthew; Braconnier, Axelle; Braga, Cynthia; Brandenburger, Timo; Brás Monteiro, Filipa; Brazzi, Luca; Breen, Dorothy; Breen, Patrick; Brickell, Kathy; Browne, Alex; Brozzi, Nicolas; Buesaquillo, Christian; Buisson, Marielle; Burhan, Erlina; Bustos, Ingrid G.; Butler, Amelia; Cabie, André; Cabral, Susana; Caceres, Eder; Cadoz, Cyril; Calligy, Kate; Calvache, Jose Andres; Camões, João; Campana, Valentine; Campbell, Paul; Canepa, Cecilia; Cantero, Mireia; Caraux-Paz, Pauline; Cárcel, Sheila; Cardellino, Chiara; Cardoso, Filipa; Cardoso, Filipe; Cardoso, Nelson; Cardoso, Sofia; Carelli, Simone; Carlier, Nicolas; Carmoi, Thierry; Carney, Gayle; Carpenter, Chloe; Carret, Marie-Christine; Carrier, François Martin; Carson, Gail; Casanova, Maire-Laure; Cascão, Mariana; Casimiro, José; Castañeda, Silvia; Castanheira, Nidyanara; Castor-Alexandre, Guylaine; Castrillón, Henry; Castro, Ivo; Catarino, Ana; Catherine, François-Xavier; Cavalin, Roberta; Cavalli, Giulio Giovanni; Cavayas, Alexandros; Ceccato, Adrian; Cervantes-Gonzalez, Minerva; Chair, Anissa; Chakveatze, Catherine; Chan, Adrienne; Chand, Meera; Chantalat Auger, Christelle; Chapplain, Jean-Marc; Chas, Julie; Chassin, Camille; Chaudary, Mobin; Chávez Iñiguez, Jonathan Samuel; Chen, Anjellica; Chen, Yih-Sharng; Cheng, Matthew Pellan; Cheret, Antoine; Chiarabini, Thibault; Chica, Julian; Chidambaram, Suresh Kumar; Chirouze, Catherine; Chiumello, Davide; Cho, Hwa Jin; Cho, Sung Min; Cho, Young-Jae; Cholley, Bernard; Chopin, Marie-Charlotte; Chua, Hiu Jian; Cidade, Jose Pedro; Cisneros Herreros, Jose Miguel; Citarella, Barbara Wanjiru; Ciullo, Anna; Clarke, Jennifer; Clohisey, Sara; Coca, Necsoi; Codan, Cassidy; Cody, Caitriona; Coelho, Alexandra; Colin, Gwenhaël; Collins, Michael; Colombo, Sebastiano Maria; Combs, Pamela; Connor, Marie; Conrad, Anne; Contreras, Sofía; Conway, Elaine; Cooke, Graham S.; Copland, Mary; Cordel, Hugues; Corley, Amanda; Cormican, Sarah; Cornelis, Sabine; Corpuz, Arianne Joy; Corvaisier, Grégory; Couffignal, Camille; Couffin-Cadiergues, Sandrine; Courtois, Roxane; Cousse, Stéphanie; Croonen, Sabine; Crowl, Gloria; Crump, Jonathan; Cruz, Claudina; Cruz Bermúdez, Juan Luis; Cruz Rojo, Jaime; Csete, Marc; Cucino, Alberto; Cullen, Caroline; Cummings, Matthew; Curley, Gerard; Curlier, Elodie; Custodio, Paula; D'Amico, Federico; D'Aragon, Frédérick; D'Ortenzio, Eric; da Silva Filipe, Ana; Da Silveira, Charlene; Dabaliz, Al-Awwab; Dagens, Andrew; Dalton, Heidi; Dalton, Jo; Daneman, Nick; Daniel, Corinne; Dankwa, Emmanuelle; Dantas, Jorge; Dantas, Vicente; de Boer, Mark; de Mendoza, Diego; De Montmollin, Etienne; de Oliveira França, Rafael Freitas; de Pinho Oliveira, Ana Isabel; De Rosa, Rosanna; de Silva, Thushan; De vries, Peter; Deacon, Jillian; Dean, David; Debard, Alexa; Debray, Marie-Pierre; DeCastro, Nathalie; Dechert, William; Deconninck, Lauren; Decours, Romain; Defous, Eve; Delacroix, Isabelle; Delaneuve, Eric; Delavigne, Karen; Delfos, Nathalie M.; Deligiannis, Ionna; Dell'Amore, Andrea; Delmas, Christelle; Delobel, Pierre; Demonchy, Elisa; Denis, Emmanuelle; Deplanque, Dominique; Depuydt, Pieter; Desai, Mehul; Descamps, Diane; Desvallée, Mathilde; Dewayanti, Santi; Diallo, Alpha; Diamantis, Sylvain; Dias, André; Diaz, Juan Jose; Diaz, Priscila; Diaz, Rodrigo; Didier, Kévin; Diehl, Jean-Luc; Dieperink, Wim; Dimet, Jérôme; Dinot, Vincent; Diop, Fara; Diouf, Alphonsine; Dishon, Yael; Djossou, Félix; Dobrita, Ana; Docherty, Annemarie B.; Dominguez, Carmen Infante; Dondorp, Arjen M; Donnelly, Christl A.; Donnelly, Maria; Donohue, Chloe; Dorival, Céline; Douglas, James Joshua; Douma, Renee; Dournon, Nathalie; Downer, Triona;

Downing, Mark; Drake, Tom; Driscoll, Aoife; Duarte Fonseca, Claudio; Dubee, Vincent; Dubos, François; Ducancelle, Alexandre; Duculan, Toni; Dudman, Susanne; Dunand, Paul; Dunning, Jake; Duplaix, Mathilde; Durante Mangoni, Emanuele; Durham III, Lucian; Dussol, Bertrand; Duthoit, Juliette; Duval, Xavier; Dyrhol-Riise, Anne Margarita; Echeverria-Villalobos, Marco; Egan, Siobhan; Eira, Carla; El Sanharawi, Mohammed; Elapavaluru, Subbarao; Elharrar, Brigitte; Ellerbroek, Jacobien; Ellis, Rachael; Eloy, Philippine; Elshazly, Tarek; Enderle, Isabelle; Engelmann, Ilka; Enouf, Vincent; Epaulard, Olivier; Escher, Martina; Esperatti, Mariano; Esperou, Hélène; Esposito-Farese, Marina; Estevão, João; Etienne, Manuel; Ettalhaoui, Nadia; Everding, Anna; Evers, Mirjam; Fabre, Isabelle; Faheem, Amna; Fahy, Arabella; Fairfield, Cameron J.; Faria, Pedro; Farooq, Ahmed; Farshait, Nataly; Fatoni, Arie Zainul; Faure, Karine; Favory, Raphaël; Fayed, Mohamed; Feely, Niamh; Fernandes, Jorge; Fernandes, Marília; Fernandes, Susana; Ferrand, François-Xavier; Ferrand Devouge, Eglantine; Ferrão, Joana; Ferraz, Mário; Ferreira, Benigno; Ferrer-Roca, Ricard; Ferriere, Nicolas; Ficko, Céline; Figueiredo-Mello, Claudia; Fiorda, Juan; Fischer, Karlee; Flament, Thomas; Flateau, Clara; Fletcher, Tom; Florio, Letizia Lucia; Flynn, Brigid; Foley, Claire; Fonseca, Tatiana; Fontanese, Nicole; Fontela, Patricia; Forsyth, Simon; Foster, Denise; Foti, Giuseppe; Fourn, Erwan; Fowler, Robert A.; Fraher, Dr Marianne; France, Dawn; Franch-Llasat, Diego; Fraser, Christophe; Fraser, John F.; Freire, Marcela Vieira; Freitas Ribeiro, Ana; Friedrich, Caren; Fritz, Ricardo; Fry, Stéphanie; Fuentes, Nora; Fukuda, Masahiro; Gaborieau, Valérie; Gaci, Rostane; Gagliardi, Massimo; Gagnard, Jean-Charles; Gagné , Nathalie; Gagneux-Brunon, Amandine; Gaiao, Sergio; Gail Skeie, Linda; Gallagher, Phil; Gallego Curto, Elena; Gamble, Carrol; Garan, Arthur; Garcia, Rebekha; García Barrio, Noelia; Garcia-Gallo, Esteban; Garot, Denis; Garrait, Valérie; Gault, Nathalie; Gavin, Aisling; Gavrilov, Anatoliy; Gaymard, Alexandre; Gebauer, Johanes; Geraud, Eva; Gerbaud Morlaes, Louis; Germano, Nuno; Ghosn, Jade; Giani, Marco; giaquinto, carlo; Gibson, Jess; Gigante, Tristan; Gilg, Morgane; Giordano, Guillermo; Girijadevi, Dinuraj; Girvan, Michelle; Gissot, Valérie; Giwangkancana, Gezy; Glikman, Daniel; Gnall, Eric; Goco, Geraldine; Goehringer, François; Goepel, Siri; Goffard, Jean-Christophe; Golob, Jonathan; Gomes, Rui; Gómez-Junyent, Joan; Gominet, Marie; Gonzalez Gonzalez, Alicia; Gorenne, Isabelle; Goubert, Laure; Goujard, Cécile; Goulenok, Tiphaine; Graafland, A; Grable, Margarite; Graf, Jeronimo; Grandin, Edward Wilson; Granier, Pascal; Grasselli, Giacomo; Grazioli, Lorenzo; Green, Christopher A.; Greenhalf, William; Greffe, Segolène; Grieco, Domenico Luca; Griffee, Matthew; Griffiths, Fiona; Grigoras, Ioana; Groenendijk, Albert; Grosse Lordemann, Anja; Gruner, Heidi; Gu, Yusing; Guarracino, Fabio; Guedj, Jérémie; Guego, Martin; Guellec, Dewi; Guerguerian, Anne-Marie; Guerreiro, Daniela; Guery, Romain; Guillaumot, Anne; Guilleminault, Laurent; Guimard, Thomas; Gutierrez, Ashley; Haber, Daniel; Hachemi, Ali; Hadri, Nadir; Hakak, Sheeba; Hall, Adam; Hall, Matthew; Halpin, Sophie; Hamer, Ansley; Hamidfar, Rebecca; Hammond, Terese; Haniffa, Rashan; Hardwick, Hayley; Harley, Kristen; Harrison, Ewen M.; Harrison, Janet; Harrison, Samuel Bernard Ekow; Hasanova, Amina; Hayat, Muhammad; Hays, Leanne; Heerman, Jan; Heggelund, Lars; Hendry, Ross; Hennessy, Martina; Henriquez, Aquiles; Hentzien, Maxime; Herekar, Fivzia; Hernandez-Montfort, Jaime; Herr, Daniel; Hershey, Andrew; Hesstvedt, Liv; Hidaya, Astarini; Higgins, Dawn; Higgins, Eibhilin; Hinton, Samuel; Hipólito-Reis, Ana; Hiraiwa, Hiroaki; Hitoto, Hikombo; Ho, Antonia Ying Wai; Hoctin, Alexandre; Hoffman, Julie; Hoffmann, Isabelle; Hoiting, Oscar; Holt, Rebecca; Holter, Jan Cato; Horby, Peter; Horcajada, Juan Pablo; Hoshino, Koji; Hoshino, Kota; Houas, Ikram;

Hough, Catherine L.; Hsu, Jimmy Ming-Yang; Hulot, Jean-Sébastien; Ijaz, Samreen; Illes, Hajnal-Gabriela; Imbert, Patrick; Inácio, Hugo; Iosifidis, Elias; Irvine, Lacey; Isgett, Sarah; Ishani, Palliya Guruge Pramodya Ishani; Isidoro, Tiago; Isnard, Margaux; Itai, Junji; Ivulich, Daniel; Iwasaki, Yudai; Jaafar, Danielle; Jaafoura, Salma; Jabot, Julien; Jackson, Clare; Jacquet, Pierre; Jamieson, Nina; Jaud-Fischer, Coline; Jaureguiberry, Stéphane; Jawad, Issrah; Jayakumar, Devachandran; Jego, Florence; Jenum, Synne; Jorge Garcia, Ruth; Joseph, Cédric; Joseph, Mark; Joshi, Swosti; Jourdain, Mercé; Jouvet, Philippe; Jung, Anna; Jung, Hanna; Juzar, Dafsah; Kafif, Ouifiya; Kaguelidou, Florentia; Kali, Sabina; Kalomoiri, Smaragdi; Kamal, Saima; Kambiya, Paul; Kandamby, Darshana; Kandel, Chris; Kant, Ravi; Kanyawati, Dyah; Karam, Isabela; Kartsonaki, Christiana; Kasugai, Daisuke; Kataria, Anant; Katz, Kevin; Kaur Johal, Simreen; Kawasaki, Tatsuya; Kay, Christy; Keating, Sean; Kelly, Andrea; Kelly, Sadie; Kennedy, Lisa; Kennedy, Ryan; Kennon, Kalynn; Kerroumi, Younes; Kestelyn, Evelyne; Khalid, Imrana; Khalid, Osama; Khalil, Antoine; Khan, Coralie; Khan, Irfan; Khanal, Sushil; Kho, Michelle E; Khoo, Saye; Khoso, Nasir; Kida, Yuri; Kiiza, Peter; Kildal, Anders Benjamin; Kim, Jae Burm; Kimmoun, Antoine; Kindgen-Milles, Detlef; Kitamura, Nobuya; Klenerman, Paul; Kloumann Bekken, Gry; Knight, Stephen; Kobbe, Robin; Kodippily, Chamira; Kohns Vasconcelos, Malte; Koirala, Sabin; Komatsu, Mamoru; Korten, Volkan; Kosgei, Caroline; Kpangon, Arsène; Krawczyk, Karolina; Kruglova, Oksana; Kumar, Ashok; Kumar, Deepali; Kumar, Mukesh; Kumar Tirupakuzhi Vijayaraghavan, Bharath; Kumar Vecham, Pavan; Kurtzman, Ethan; Kusumastuti, Neurinda Permata; Kutsogiannis, Demetrios; Kutsyna, Galyna; Kyriakoulis, Konstantinos; L'Her, Erwan; Lachatre, Marie; Lacoste, Marie; Laffey, John G; Lagrange, Marie; Laine, Fabrice; Lairez, Olivier; Lalueza Blanco, Antonio; Lambert, Marc; Lamontagne, François; Langelot-Richard, Marie; Langlois, Vincent; Lantang, Eka Yudha; Lanza, Marina; Laouénan, Cédric; Laribi, Samira; Lariviere, Delphine; Lasry, Stéphane; Launay, Odile; Laureillard, Didier; Lavie-Badie, Yoan; Law, Andrew; Lawrence, Cassie; Le, Minh; Le Bihan, Clément; Le Bris, Cyril; Le Falher, Georges; Le Fevre, Lucie; Le Hingrat, Quentin; Le Maréchal, Marion; Le Mestre, Soizic; Le Moal, Gwenaël; Le Moing, Vincent; Le Nagard, Hervé; Le Turnier, Paul; Leal, Ema; Leal Santos, Marta; Lee, James; Lee, Su  Hwan; Lee, Todd C.; Leeming, Gary; Lefebvre, Bénédicte; Lefebvre, Laurent; Lefevre, Benjamin; LeGac, Sylvie; Lelievre, Jean-Daniel; Lellouche, François; Lemaignen, Adrien; Lemee, Véronique; Lemeur, Anthony; Lemmink, Gretchen; León, Rafael; Leone, Marc; Leone, Michela; Lepiller, Quentin; Lescure, François-Xavier; Lesens, Olivier; Lesouhaitier, Mathieu; Levy, Bruno; Levy, Yves; Levy-Marchal, Claire; Li Bassi, Gianluigi; Liang, Janet; Liaquat, Ali; Liegeon, Geoffrey; Lim, Wei Shen; Lima, Chantre; Lina, Bruno; Lind, Andreas; Lingas, Guillaume; Link, Linda; Lion-Daolio, Sylvie; Liu, Keibun; Livrozet, Marine; Lizotte, Patrica; Loforte, Antonio; Lolong, Navy; Lopes, Diogo; Lopez-Colon, Dalia; Loschner, Anthony L.; Lotz, Goesta; Loubet, Paul; Loufti, Bouchra; Louis, Guillame; Lourenco, Silvia; Lucet, Jean Christophe; Lumbreras Bermejo, Carlos; Luna, Carlos M.; Lungu, Olguta; Luong, Liem; Luque, Nestor; Luton, Dominique; Lwin, Nilar; Lyons, Ruth; Maasikas, Olavi; Mabiala, Oryane; MacDonald, Samual; Machado, Moïse; Macheda, Gabriel; Macias Sanchez, Juan; Madhok, Jai; Madiha, Hashmi; Madiha, Hashmi; Maestro de la Calle, Guillermo; Mahieu, Rafael; Mahy, Sophie; Maia, Ana Raquel; Maier, Lars Siegfried; Maillet, Mylène; Maitre, Thomas; Malfertheiner, Maximilian; Malik, Nadia; Maltez, Fernando; Malvy, Denis; Mambert, Marina; Manda, Victoria; Mandei, Jose M.; Mandelbrot, Laurent; Mankikian, Julie; Manning, Edmund; Manuel, Aldric; Maria Sant`Ana Malaque, Ceila; Marino, Daniel;

Marino, Flávio; Mariz, Carolline de Araújo; Markowicz, Samuel; Maroun Eid, Charbel; Marques, Ana; Marquis, Catherine; Marsh, Brian; Marsh, Laura; Marshall, John; Martelli, Celina Turchi; Martin, Emily; Martin-Blondel, Guillaume; Martin-Loeches, Ignacio; Martin-Quiros, Alejandro; Martinelli, Alessandra; Martinot, Martin; Martins, Ana; Martins, João; Martins, Nuno; Martins Rego, Caroline; Martucci, Gennaro; Martynenko, Olga; Marwali, Eva Miranda; Masa Jimenez, Juan Fernado; Maslove, David; Mason, Sabina; Mat Nor, Basri; Matan, Moshe; Mathieu, Daniel; Mattei, Mathieu; Matulevics, Romans; Maulin, Laurence; May, Jennifer; Maynar, Javier; Mazzoni, Thierry; Mc Evoy, Natalie; McArthur, Colin; McBride, Angela; McCarthy, Aine; McCarthy, Anne; McCloskey, Colin; McConnochie, Rachael; McDermott, Sherry; McDonald, Sarah; McElwee, Samuel; McGeer, Allison; McGuinness, Niki; McKay, Chris; McKeown, Johnny; McLean, Kenneth A.; McNicholas, Bairbre; Meaney, Edel; Mear-Passard, Cécile; Mechlin, Maggie; Meher, Maqsood; Mehkri, Omar; Mele, Ferruccio; Melo, Luis; Memon, Kashif; Mendes, Joao Joao; Menkiti, Ogechukwu; Menon, Kusum; Mentré, France; Mentzer, Alexander J.; Mercier, Emmanuelle; Mercier, Noémie; Merckx, Antoine; Mergeay-Fabre, Mayka; Mergler, Blake; Merson, Laura; Mesquita, António; Meybeck, Agnès; Meyer, Dan; Meynert, Alison M.; Meysonnier, Vanina; Meziane, Amina; Mezidi, Mehdi; Michelagnoli, Giuliano; Michelanglei, Céline; Michelet, Isabelle; Mihelis, Efstathia; Mihnovitš, Vladislav; Moin, Asma; Molina, David; Molinos, Elena; Molloy, Alex; Mone, Mary; Monteiro, Agostinho; Montes, Claudia; Montrucchio, Giorgia; Moore, Sarah; Moore, Shona C.; Morales Cely, Lina; Moro, Lucia; Motherway, Catherine; Motos, Ana; Mouquet, Hugo; Mouton Perrot, Clara; Moyet, Julien; Mullaert, Jimmy; Müller, Fredrik; Müller, Karl Erik; Muneeb, Syed; Murris, Marlène; Murthy, Srinivas; Myrodia, Dimitra Melia; Nagpal, Dave, Nagrebetsky, Alex; Narasimhan, Mangala; Nasim Khan, Rashid; Natanel, Hans; Neant, Nadège; Neb, Holger; Neto, Raul; Neumann, Emily; Neves, Bernardo; Ng, Pauline Yeung; Ng, Wing Yiu; Nghi, Anthony; Nguyen, Duc; Ni Choileain, Orna; Nichol, Alistair; Nicholson, Meghan; Nitayavardhana, Prompak; Nonas, Stephanie; Noret, Marion; Norman, Lisa; Notari, Alessandra; Noursadeghi, Mahdad; Nowicka, Karolina; Nseir, Saad; Nunez, Jose I; Nurnaningsih, Nurnaningsih; Nyamankolly, Elsa; O'Donnell, Max; O'Hearn, Katie; Occhipinti, Giovanna; Ogston, Tawnya; Ogura, Takayuki; Oh, Tak-Hyuk; Ohshimo, Shinichiro; Oinam, Budhacharan Singh; Oldakowska, Agnieszka; Oliveira, João; Oliveira, Larissa; Olliaro, Piero L.; Ong, David S.Y.; Oosthuyzen, Wilna; Opavsky, Anne; Openshaw, Peter; Orakzai, Saijad; Orozco-Chamorro, Claudia Milena; Ortoleva, Jamel; Osatnik, Javier; Ouamara, Nadia; Ouissa, Rachida; Owyang, Clark; Oziol, Eric; Pabasara, H M Upulee; Pagadoy, Maïder; Pages, Justine; Palacios, Amanda; Palacios, Mario; Palmarini, Massimo; Panarello, Giovanna; Panda, Prasan Kumar; Paneru, Hem; Panigada, Mauro; Pansu, Nathalie; Papadopoulos, Aurélie; Parke, Rachael; Parker, Melissa; Parra, Briseida; Parrini, Vieri; Pasha, Taha; Pasquier, Jérémie; Pastene, Bruno; Patauner, Fabian; Patel, Junaid; Pathmanathan, Mohan Dass; Patrão, Luís; Patricio, Patricia; Patrier, Juliette; Patterson, Lisa; Pattnaik, Rajyabardhan; Paul, Christelle; Paul, Mical; Paulos, Jorge; Paxton, William A.; Payen, Jean-François; Peariasamy, Kalaiarasu; Pearse, India; Pedrera Jiménez, Miguel; Peek, Giles; Peelman, Florent; Peiffer-Smadja, Nathan; Peigne, Vincent; Pejkovska, Mare; Pelosi, Paolo; Peltan, Ithan D.; Pereira, Rui; Perez, Daniel; Periel, Luis; Perpoint, Thomas; Pesenti, Antonio; Pestre, Vincent; Petroušová, Lenka; Petrov-Sanchez, Ventzislava; Pettersen, Frank Olav; Peytavin, Gilles; Pharand, Scott; Piagnerelli, Michael; Picard, Walter; Picone, Olivier; Piero, Maria de; Pierobon, Carola; Pimentel, Carlos; Pinilla, Ana; Pinto, Raquel; Pironneau, Isabelle; Piroth, Lionel; Pius, Riinu;

Piva, Simone; Plantier, Laurent; Plotkin, Daniel; Poissy, Julien; Pokeerbux, Ryadh; Pokorska-Śpiewak, Maria; Poli, Sergio; Pollakis, Georgios; Ponscarme, Diane; Popielska, Jolanta; Post, Andra-Maris; Postma, Douwe F.; Povoa, Pedro; Póvoas, Diana; Powis, Jeff; Prapa, Sofia; Preau, Sébastien; Prebensen, Christian; Preiser, Jean-Charles; Prinssen, Anton; Pritchard, Mark; Priyadarshani, Gamage Dona Dilanthi; Proença, Lúcia; Puéchal, Oriane; Pujo Semedi, Bambang; Pulicken, Mathew; Purcell, Gregory; Quesada, Luisa; Quinones-Cardona, Vilmaris; Quirós González, Víctor; Quist-Paulsen, Else; Quraishi, Mohammed; Rabaud, Christian; Rafael, Aldo; Rafiq, Marie; Rahutullah, Arsalan; Rainieri, Fernando; Ralib, Azrina; Ramakrishnan, Nagarajan; Rammaert, Blandine; Rana, Asim; Rapp, Christophe; Rashan, Aasiyah; Rashan, Thalha; Rasmin, Menaldi; Ratsep, Indrek; Rau, Cornelius; Raza, Ali; Real, Andre; Rebaudet, Stanislas; Redl, Sarah; Reeve, Brenda; Rehan, Ali; Rehman, Attaur; Reid, Liadain; Reikvam, Dag Henrik; Reis, Renato; Rello, Jordi; Remppis, Jonathan; Remy, Martine; Ren, Hongru; Renk, Hanna; Resende, Liliana; Resseguier, Anne-Sophie; Revest, Matthieu; Rewa, Oleksa; Reyes, Luis F.; Reyes, Tiago; Ribeiro, Maria Ines; Richardson, David; Richardson, Denise; Richier, Laurent; Riera, Jordi; Rios, Ana Lúcia; Rishu, Asgar; Rispal, Patrick; Risso, Karine; Rivera Nuñez, Maria Angelica; Rizer, Nicholas; Robba, Chiara; Roberto, André; Roberts, Stephanie; Robertson, David L.; Robineau, Olivier; Roche-Campo, Ferran; Rodari, Paola; Rodeia, Simão; Rodriguez Abreu, Julia; Roger, Claire; Roger, Pierre-Marie; Roilides, Emmanuel; Rojek, Amanda; Romaru, Juliette; Roncon-Albuquerque Jr, Roberto; Roriz, Mélanie; Rosa-Calatrava, Manuel; Rose, Michael; Rosenberger, Dorothea; Rossanese, Andrea; Rossetti, Matteo; Rossignol, Bénédicte; Rossignol, Patrick; Rousset, Stella; Roy, Carine; Roze, Benoît; Rusmawatiningtyas, Desy; Russell, Clark D.; Ryckaert, Steffi; Rygh Holten, Aleksander; Saba, Isabela; Sadaf, Sairah; Sadat, Musharaf; Sahraei, Valla; Saint-Gilles, Maximilien; Sakiyalak, Pranya; Salahuddin, Nawal; Salazar, Leonardo; Sales, Gabriele; Salgado, Juan; Sallaberry, Stéphane; Salmon Gandonniere , Charlotte; Salvator, Hélène; Sanchez, Angel; Sanchez, Olivier; Sancho-Shimizu, Vanessa; Sandhu, Gyan; Sandrine, Pierre-François; Sandulescu, Oana; Santana, Sergio Ruiz; Santos, Marlene; Sarfo-Mensah, Shirley; Sarmiento, Iam Claire E.; Sarton, Benjamine; Satyapriya, Sree; Satyawati, Rumaisah; Saviciute, Egle; Savvidou, Parthena; Scarsbrook, Joshua; Schaffer, Justin; Schermer, Tjard; Scherpereel, Arnaud; Schneider, Marion; Schroll, Stephan; Scott, Janet T.; Scott-Brown, James; Sedillot, Nicholas; Seefeldt, Cassandra; Seitz, Tamara; Selvanayagam, Jaganathan; Semaille, Caroline; Semple, Malcolm G.; Senneville, Eric; Sepulveda, Claudia; Sequeira, Filipa; Sequeira, Tânia; Serrano Balazote, Pablo; Shadowitz, Ellen; Shamsah, Mohammad; Sharma, Pratima; Shaw, Catherine A.; Shaw, Victoria; Sheharyar, Ashraf; Shi, Haixia; Shiekh, Mohiuddin; Shime, Nobuaki; Shimizu, Hiroaki; Shimizu, Keiki; Shimizu, Naoki; Shrapnel, Sally; Shum, Hoi Ping; Si Mohammed, Nassima; Sibiude, Jeanne; Siddiqui, Atif; Sigfrid, Louise; Sillaots, Piret; Silva, Catarina; Silva, Maria Joao; Silva, Rogério; Sim Lim Heng, Benedict; Sin, Wai Ching; Sitompul, Pompini Agustina; Skogen, Vegard; Smith, Sue; Smood, Benjamin; Smyth, Michelle; Snacken, Morgane; So, Dominic; Solis, Monserrat; Solomon, Joshua; Solomon, Tom; Somers, Emily; Sommet, Agnès; Song, Myung Jin; Song, Rima; Song, Tae; Sonntagbauer, Michael; Sotto, Alberto; Soum, Edouard; Sousa, Ana Chora; Sousa, Marta; Sousa Uva, Maria; Souza-Dantas, Vicente; Sperry, Alexandra; Sri Darshana, B. P. Sanka Ruwan; Sriskandan, Shiranee; Stabler, Sarah; Stecher, Stepanie-Susanne; Stienstra, Ymkje; Stiksrud, Birgitte; Streinu-Cercel, Adrian; Streinu-Cercel, Anca; Strudwick, Samantha; Stuart, Ami; Stuart, David; Suen, Gabriel; Suen,

Jacky Y.; Sultana, Asfia; Summers, Charlotte; Surovcova, Magdalena; Swanson, Ashleigh; Syrigos, Konstantinos; Sztajnbok, Jaques; Szuldrzynski, Konstanty; Tabrizi, Shirin; Taccone, Fabio; Tagherset, Lysa; Talarek, Ewa; Taleb, Sara; Talsma, Jelmer; Tampubolon, Maria Lawrensia; Tan, Le Van; Tanaka, Hiro; Tanaka, Taku; Taniguchi, Hayato; Tanveer, Hussain; Tardivon, Coralie; Tattevin, Pierre; Tedder, Richard S.; Teixeira, João; Tejada, Sofia; Tellier, Marie-Capucine; Teotonio, Vanessa; Téoulé, François; Terpstra, Pleun; Terrier, Olivier; Terzi, Nicolas; Tessier-Grenier, Hubert; Thibault, Vincent; Thiberville, Simon-Djamel; Thill, Benoît; Thompson, Pat; Thompson, Shaun; Thomson, David; Thomson, Emma C.; Thuy, Duong Bich; Thwaites, Ryan S.; Tieroshyn, Vadim; Timsit, Jean-François; Tissot, Noémie; Toki, Maria; Tolppa, Timo; Tolwani, Ashita; Tonby, Kristian; Torres, Antoni; Torres, Margarida; Torres Santos-Olmo, Rosario Maria; Torres- Zevallos, Hernando; Townsend, Joan; Treoux, Théo; Trieu, Huynh Trung; Tromeur, Cécile; Trontzas, Ioannis; Troost, Jonathan; Trouillon, Tiffany; Truong, Jeanne; Tual, Christelle; Tubiana, Sarah; Tuite, Helen; Turmel, Jean-Marie; Turtle, Lance C.W.; Tveita, Anders; Twardowski, Pawel; Uchiyama, Makoto; Udayanga, PG Ishara; Uribe, Alberto; Usman, Asad; Val-Flores, Luís; Valle, Ana Luiza; Valran, Amélie; Van De Velde, Stijn; Van der Feltz, Machteld; Van Der Vekens, Nicky; Van der Voort, Peter; Van Der Werf, Sylvie; van Gulik, Laura; Van Hattem, Jarne; van Lelyveld, Steven; van Netten, Carolien; van Twillert, G; Vanel, Noémie; Vanoverschelde, Henk; Vauchy, Charline; Veislinger, Aurélie; Ventura, Sara; Verbon, Annelies; Vercaemst, Leen; Vidal, José Ernesto; Vieira, César; Villanueva, Joy Ann; Villar, Judit; Villeneuve, Pierre-Marc; Villoldo, Andrea; Vinh Chau, Nguyen Van; Visseaux, Benoit; Visser, Hannah; Vitiello, Chiara; Vuorinen, Aapeli; Vuotto, Fanny; Walter, Alicia; Wan Muhd Shukeri, Wan Fadzlina; Wang, Chih-Hsien; Wei, Jia; Weil, Katharina; Wesselius, Sanne; Wham, Murray; Whelan, Bryan; White, Nicole; Wicky, Paul Henri; Wiedemann, Aurélie; Wijaya, Surya Oto; Wille, Keith; Willems, Suzette; Williams, Virginie; Wils, Evert-Jan; Xynogalas, Ioannis; Yacoub, Sophie; Yamazaki, Masaki; Yazdanpanah, Yazdan; Yelnik, Cécile; Yerkovich, Stephanie; Yokoyama, Toshiki; Yonis, Hodane; Yuliarto, Saptadi; Zaaqoq, Akram; Zabbe, Marion; Zacharowski, Kai; Zahran, Maram; Zambon, Maria; Zambrano, Miguel; Zanella, Alberto; Zawadka, Konrad; Zayyad, Hiba; Zoufaly, Alexander; Zucman, David.
