## Supplemental table S1 for "Ten months of temporal variation in the clinical journey of hospitalised patients with COVID-19: an observational cohort"

| Variable | Value | Total | March | April | May | June | July | August | September | October | November | December | 2021 |
|---|---|---|---|---|---|---|---|---|---|---|---|---|---|
| Sex | Female | 60977 (42.8) | 10709 (39.5) | 18102 (42.8) | 5732 (46.6) | 2366 (44.3) | 1200 (42.7) | 865 (39) | 2233 (42.4) | 5912 (42.8) | 6642 (43.8) | 5958 (45.1) | 1258 (41.4) |
| | Male | 81325 (57.1) | 16347 (60.3) | 24092 (57) | 6565 (53.3) | 2972 (55.6) | 1608 (57.2) | 1350 (60.9) | 3015 (57.3) | 7887 (57.1) | 8485 (56) | 7229 (54.7) | 1775 (58.5) |
| | Unknown | 238 (0.167) | 52 (0.192) | 73 (0.173) | 14 (0.114) | 4 (0.0749) | 3 (0.107) | 3 (0.135) | 17 (0.323) | 23 (0.166) | 28 (0.185) | 18 (0.136) | 3 (0.0988) |
| Age group | 0-19 | 2730 (1.92) | 581 (2.14) | 563 (1.33) | 319 (2.59) | 129 (2.41) | 68 (2.42) | 59 (2.66) | 113 (2.15) | 258 (1.87) | 310 (2.05) | 297 (2.25) | 33 (1.09) |
| | 20-39 | 9546 (6.7) | 1701 (6.27) | 2423 (5.73) | 759 (6.17) | 478 (8.95) | 334 (11.9) | 261 (11.8) | 550 (10.4) | 890 (6.44) | 964 (6.36) | 942 (7.13) | 244 (8.04) |
| | 40-59 | 31368 (22) | 5756 (21.2) | 9223 (21.8) | 2308 (18.7) | 1167 (21.8) | 720 (25.6) | 658 (29.7) | 1280 (24.3) | 2904 (21) | 3273 (21.6) | 3110 (23.6) | 969 (31.9) |
| | 60-69 | 23431 (16.4) | 4581 (16.9) | 6885 (16.3) | 1769 (14.4) | 872 (16.3) | 459 (16.3) | 361 (16.3) | 897 (17) | 2303 (16.7) | 2590 (17.1) | 2098 (15.9) | 616 (20.3) |
| | 70-79 | 30541 (21.4) | 5854 (21.6) | 8988 (21.3) | 2516 (20.4) | 1034 (19.4) | 473 (16.8) | 344 (15.5) | 1095 (20.8) | 3356 (24.3) | 3497 (23.1) | 2744 (20.8) | 640 (21.1) |
| | 80+ | 42092 (29.5) | 7099 (26.2) | 13967 (33) | 4555 (37) | 1470 (27.5) | 548 (19.5) | 341 (15.4) | 1186 (22.5) | 4017 (29.1) | 4439 (29.3) | 3949 (29.9) | 521 (17.2) |
| | Unknown | 2832 (1.99) | 1536 (5.67) | 218 (0.516) | 85 (0.69) | 192 (3.59) | 209 (7.44) | 194 (8.75) | 144 (2.74) | 94 (0.68) | 82 (0.541) | 65 (0.492) | 13 (0.428) |
| Nosocomial infection | No | 121910 (85.5) | 23620 (87.1) | 36942 (87.4) | 10158 (82.5) | 4509 (84.4) | 2417 (86) | 1959 (88.3) | 4486 (85.2) | 11417 (82.6) | 12587 (83.1) | 10779 (81.6) | 3036 (100) |
| | Yes | 11695 (8.2) | 2575 (9.5) | 4045 (9.57) | 1209 (9.82) | 346 (6.48) | 138 (4.91) | 62 (2.8) | 213 (4.05) | 910 (6.58) | 1092 (7.21) | 1105 (8.37) | 0 (0) |
| | Unknown | 8935 (6.27) | 913 (3.37) | 1280 (3.03) | 944 (7.67) | 487 (9.12) | 256 (9.11) | 197 (8.88) | 566 (10.8) | 1495 (10.8) | 1476 (9.74) | 1321 (10) | 0 (0) |
| Pregnancy | Yes | 1030 (0.723) | 176 (0.649) | 200 (0.473) | 67 (0.544) | 41 (0.768) | 42 (1.49) | 37 (1.67) | 68 (1.29) | 124 (0.897) | 119 (0.785) | 130 (0.984) | 26 (0.856) |
| | No | 139630 (98) | 26545 (97.9) | 41524 (98.2) | 12111 (98.4) | 5252 (98.3) | 2754 (98) | 2145 (96.7) | 5095 (96.8) | 13518 (97.8) | 14829 (97.8) | 12901 (97.7) | 2956 (97.4) |
| | Unknown | 1880 (1.32) | 387 (1.43) | 543 (1.28) | 133 (1.08) | 49 (0.917) | 15 (0.534) | 36 (1.62) | 102 (1.94) | 180 (1.3) | 207 (1.37) | 174 (1.32) | 54 (1.78) |

Table S1

| Variable | Value | Total | March | April | May | June | July | August | September | October | November | December | 2021 |
|---|---|---|---|---|---|---|---|---|---|---|---|---|---|
| Abdominal pain | Yes | 13500 (9.47) | 2275 (8.39) | 3877 (9.17) | 1486 (12.1) | 601 (11.3) | 284 (10.1) | 221 (9.96) | 634 (12) | 1344 (9.72) | 1435 (9.47) | 1137 (8.61) | 206 (6.79) |
| | No | 109285 (76.7) | 20664 (76.2) | 31543 (74.6) | 9136 (74.2) | 4187 (78.4) | 2336 (83.1) | 1834 (82.7) | 4156 (78.9) | 10954 (79.3) | 11816 (78) | 10255 (77.7) | 2404 (79.2) |
| | Unknown | 19755 (13.9) | 4169 (15.4) | 6847 (16.2) | 1689 (13.7) | 554 (10.4) | 191 (6.79) | 163 (7.35) | 475 (9.02) | 1524 (11) | 1904 (12.6) | 1813 (13.7) | 426 (14) |
| Ageusia | Yes | 7103 (4.98) | 429 (1.58) | 1308 (3.09) | 453 (3.68) | 263 (4.92) | 163 (5.8) | 150 (6.76) | 455 (8.64) | 1235 (8.94) | 1317 (8.69) | 985 (7.46) | 345 (11.4) |
| | No | 71571 (50.2) | 7142 (26.3) | 16089 (38.1) | 7574 (61.5) | 3582 (67.1) | 1871 (66.6) | 1495 (67.4) | 3525 (67) | 9399 (68) | 10082 (66.5) | 8879 (67.2) | 1933 (63.7) |
| | Unknown | 63866 (44.8) | 19537 (72.1) | 24870 (58.8) | 4284 (34.8) | 1497 (28) | 777 (27.6) | 573 (25.8) | 1285 (24.4) | 3188 (23.1) | 3756 (24.8) | 3341 (25.3) | 758 (25) |
| Anosmia | Yes | 5550 (3.89) | 318 (1.17) | 901 (2.13) | 336 (2.73) | 198 (3.71) | 163 (5.8) | 145 (6.54) | 385 (7.31) | 997 (7.21) | 1049 (6.92) | 789 (5.98) | 269 (8.86) |
| | No | 74490 (52.3) | 7471 (27.6) | 17036 (40.3) | 7883 (64) | 3671 (68.7) | 1892 (67.3) | 1521 (68.6) | 3663 (69.6) | 9743 (70.5) | 10404 (68.7) | 9186 (69.6) | 2020 (66.5) |
| | Unknown | 62500 (43.8) | 19319 (71.3) | 24330 (57.6) | 4092 (33.2) | 1473 (27.6) | 756 (26.9) | 552 (24.9) | 1217 (23.1) | 3082 (22.3) | 3702 (24.4) | 3230 (24.5) | 747 (24.6) |
| Bleeding | Yes | 2123 (1.49) | 330 (1.22) | 622 (1.47) | 289 (2.35) | 109 (2.04) | 47 (1.67) | 31 (1.4) | 74 (1.41) | 191 (1.38) | 173 (1.14) | 227 (1.72) | 30 (0.988) |
| | No | 119729 (84) | 22559 (83.2) | 34554 (81.8) | 10219 (83) | 4625 (86.6) | 2559 (91) | 2021 (91.1) | 4666 (88.6) | 12003 (86.8) | 12972 (85.6) | 10995 (83.3) | 2556 (84.2) |
| | Unknown | 20688 (14.5) | 4219 (15.6) | 7091 (16.8) | 1803 (14.6) | 608 (11.4) | 205 (7.29) | 166 (7.48) | 525 (9.97) | 1628 (11.8) | 2010 (13.3) | 1983 (15) | 450 (14.8) |
| Confusion | Yes | 31604 (22.2) | 5260 (19.4) | 10789 (25.5) | 3532 (28.7) | 1035 (19.4) | 423 (15) | 283 (12.8) | 876 (16.6) | 2936 (21.2) | 3230 (21.3) | 2803 (21.2) | 437 (14.4) |
| | No | 96213 (67.5) | 18714 (69) | 26495 (62.7) | 7617 (61.9) | 3886 (72.7) | 2244 (79.8) | 1803 (81.3) | 4011 (76.2) | 9693 (70.1) | 10500 (69.3) | 8993 (68.1) | 2257 (74.3) |
| | Unknown | 14723 (10.3) | 3134 (11.6) | 4983 (11.8) | 1162 (9.44) | 421 (7.88) | 144 (5.12) | 132 (5.95) | 378 (7.18) | 1193 (8.63) | 1425 (9.4) | 1409 (10.7) | 342 (11.3) |
| Conjunctivitis | Yes | 564 (0.396) | 143 (0.528) | 154 (0.364) | 69 (0.56) | 27 (0.505) | 17 (0.605) | 10 (0.451) | 17 (0.323) | 37 (0.268) | 44 (0.29) | 35 (0.265) | 11 (0.362) |
| | No | 114954 (80.6) | 21287 (78.5) | 32966 (78) | 9916 (80.5) | 4584 (85.8) | 2528 (89.9) | 1972 (88.9) | 4561 (86.6) | 11666 (84.4) | 12492 (82.4) | 10558 (80) | 2424 (79.8) |
| | Unknown | 27022 (19) | 5678 (20.9) | 9147 (21.6) | 2326 (18.9) | 731 (13.7) | 266 (9.46) | 236 (10.6) | 687 (13) | 2119 (15.3) | 2619 (17.3) | 2612 (19.8) | 601 (19.8) |
| Cough | Yes | 89438 (62.7) | 20005 (73.8) | 28390 (67.2) | 6343 (51.5) | 2603 (48.7) | 1473 (52.4) | 1226 (55.3) | 3135 (59.5) | 8221 (59.5) | 8625 (56.9) | 7197 (54.5) | 2220 (73.1) |
| | No | 44371 (31.1) | 5599 (20.7) | 11638 (27.5) | 5273 (42.8) | 2497 (46.7) | 1235 (43.9) | 892 (40.2) | 1904 (36.2) | 4828 (34.9) | 5429 (35.8) | 4489 (34) | 587 (19.3) |
| | Unknown | 8731 (6.13) | 1504 (5.55) | 2239 (5.3) | 695 (5.65) | 242 (4.53) | 103 (3.66) | 100 (4.51) | 226 (4.29) | 773 (5.59) | 1101 (7.26) | 1519 (11.5) | 229 (7.54) |
| Diarrhoea | Yes | 22953 (16.1) | 4861 (17.9) | 7346 (17.4) | 1767 (14.4) | 695 (13) | 390 (13.9) | 315 (14.2) | 859 (16.3) | 2100 (15.2) | 2199 (14.5) | 1843 (14) | 578 (19) |
| | No | 102851 (72.2) | 18781 (69.3) | 29264 (69.2) | 9083 (73.8) | 4171 (78.1) | 2266 (80.6) | 1762 (79.4) | 3992 (75.8) | 10400 (75.2) | 11284 (74.5) | 9743 (73.8) | 2105 (69.3) |
| | Unknown | 16736 (11.7) | 3466 (12.8) | 5657 (13.4) | 1461 (11.9) | 476 (8.91) | 155 (5.51) | 141 (6.36) | 414 (7.86) | 1322 (9.56) | 1672 (11) | 1619 (12.3) | 353 (11.6) |
| Ear pain | Yes | 489 (0.343) | 133 (0.491) | 144 (0.341) | 34 (0.276) | 8 (0.15) | 9 (0.32) | 4 (0.18) | 28 (0.532) | 49 (0.355) | 38 (0.251) | 37 (0.28) | 5 (0.165) |
| | No | 96656 (67.8) | 16889 (62.3) | 28371 (67.1) | 8009 (65.1) | 3334 (62.4) | 1416 (50.4) | 992 (44.7) | 3717 (70.6) | 10493 (75.9) | 11337 (74.8) | 9831 (74.4) | 2267 (74.7) |
| | Unknown | 45395 (31.8) | 10086 (37.2) | 13752 (32.5) | 4268 (34.7) | 2000 (37.4) | 1386 (49.3) | 1222 (55.1) | 1520 (28.9) | 3280 (23.7) | 3780 (24.9) | 3337 (25.3) | 764 (25.2) |
| Fatigue | Yes | 54263 (38.1) | 10917 (40.3) | 16521 (39.1) | 4450 (36.1) | 1695 (31.7) | 969 (34.5) | 771 (34.8) | 1999 (38) | 5269 (38.1) | 5616 (37.1) | 4630 (35.1) | 1426 (47) |
| | No | 66553 (46.7) | 11404 (42.1) | 18301 (43.3) | 6006 (48.8) | 3046 (57) | 1606 (57.1) | 1260 (56.8) | 2697 (51.2) | 6939 (50.2) | 7499 (49.5) | 6589 (49.9) | 1206 (39.7) |
| | Unknown | 21724 (15.2) | 4787 (17.7) | 7445 (17.6) | 1855 (15.1) | 601 (11.3) | 236 (8.4) | 187 (8.43) | 569 (10.8) | 1614 (11.7) | 2040 (13.5) | 1986 (15) | 404 (13.3) |
| Fever | Yes | 86494 (60.7) | 19880 (73.3) | 28123 (66.5) | 6496 (52.8) | 2591 (48.5) | 1489 (53) | 1289 (58.1) | 3066 (58.2) | 7531 (54.5) | 7725 (51) | 6475 (49) | 1829 (60.2) |
| | No | 49850 (35) | 6234 (23) | 12495 (29.6) | 5328 (43.3) | 2555 (47.8) | 1233 (43.9) | 863 (38.9) | 2026 (38.5) | 5639 (40.8) | 6528 (43.1) | 5928 (44.9) | 1021 (33.6) |
| | Unknown | 6196 (4.35) | 994 (3.67) | 1649 (3.9) | 487 (3.96) | 196 (3.67) | 89 (3.17) | 66 (2.98) | 173 (3.29) | 652 (4.72) | 902 (5.95) | 802 (6.07) | 186 (6.13) |
| Headache | Yes | 13754 (9.65) | 3041 (11.2) | 3984 (9.43) | 1058 (8.59) | 468 (8.76) | 311 (11.1) | 273 (12.3) | 583 (11.1) | 1324 (9.58) | 1333 (8.8) | 1049 (7.94) | 330 (10.9) |
| | No | 100776 (70.7) | 18053 (66.6) | 28665 (67.8) | 8828 (71.7) | 4093 (76.6) | 2219 (78.9) | 1704 (76.8) | 3978 (75.6) | 10326 (74.7) | 11140 (73.5) | 9639 (73) | 2131 (70.2) |
| | Unknown | 28010 (19.7) | 6014 (22.2) | 9618 (22.8) | 2425 (19.7) | 781 (14.6) | 281 (10) | 241 (10.9) | 704 (13.4) | 2172 (15.7) | 2682 (17.7) | 2517 (19.1) | 575 (18.9) |
| Lymphadenopathy | Yes | 783 (0.549) | 155 (0.572) | 217 (0.513) | 113 (0.918) | 49 (0.917) | 17 (0.605) | 15 (0.676) | 26 (0.494) | 64 (0.463) | 67 (0.442) | 51 (0.386) | 9 (0.296) |
| | No | 113956 (79.9) | 19907 (73.4) | 33049 (78.2) | 9918 (80.6) | 4563 (85.4) | 2533 (90.1) | 1970 (88.8) | 4606 (87.5) | 11680 (84.5) | 12561 (82.9) | 10684 (80.9) | 2485 (81.9) |
| | Unknown | 27801 (19.5) | 7046 (26) | 9001 (21.3) | 2280 (18.5) | 730 (13.7) | 261 (9.28) | 233 (10.5) | 633 (12) | 2078 (15) | 2527 (16.7) | 2470 (18.7) | 542 (17.9) |
| Myalgia | Yes | 21482 (15.1) | 5198 (19.2) | 6573 (15.6) | 1487 (12.1) | 706 (13.2) | 488 (17.4) | 373 (16.8) | 844 (16) | 1931 (14) | 1812 (12) | 1509 (11.4) | 561 (18.5) |
| | No | 92380 (64.8) | 15878 (58.6) | 25896 (61.3) | 8309 (67.5) | 3840 (71.9) | 2033 (72.3) | 1609 (72.5) | 3679 (69.9) | 9701 (70.2) | 10543 (69.6) | 9010 (68.2) | 1882 (62) |
| | Unknown | 28678 (20.1) | 6032 (22.3) | 9798 (23.2) | 2515 (20.4) | 796 (14.9) | 290 (10.3) | 236 (10.6) | 742 (14.1) | 2190 (15.8) | 2800 (18.5) | 2686 (20.3) | 593 (19.5) |
| Rash | Yes | 1744 (1.22) | 315 (1.16) | 488 (1.15) | 235 (1.91) | 80 (1.5) | 32 (1.14) | 20 (0.902) | 68 (1.29) | 149 (1.08) | 167 (1.1) | 159 (1.2) | 31 (1.02) |
| | No | 114379 (80.2) | 20197 (74.5) | 33267 (78.7) | 9926 (80.6) | 4587 (85.9) | 2544 (90.5) | 1982 (89.4) | 4584 (87.1) | 11668 (84.4) | 12543 (82.8) | 10625 (80.5) | 2456 (80.9) |
| | Unknown | 26417 (18.5) | 6596 (24.3) | 8512 (20.1) | 2150 (17.5) | 675 (12.6) | 235 (8.36) | 216 (9.74) | 613 (11.6) | 2005 (14.5) | 2445 (16.1) | 2421 (18.3) | 549 (18.1) |
| Runny nose | Yes | 3745 (2.63) | 1303 (4.81) | 1101 (2.6) | 257 (2.09) | 150 (2.81) | 94 (3.34) | 58 (2.61) | 119 (2.26) | 259 (1.87) | 183 (1.21) | 180 (1.36) | 41 (1.35) |
| | No | 107192 (75.2) | 19065 (70.3) | 30549 (72.3) | 9343 (75.9) | 4348 (81.4) | 2404 (85.5) | 1891 (85.3) | 4353 (82.7) | 11052 (80) | 11875 (78.4) | 10039 (76) | 2273 (74.9) |
| | Unknown | 31603 (22.2) | 6740 (24.9) | 10617 (25.1) | 2711 (22) | 844 (15.8) | 313 (11.1) | 269 (12.1) | 793 (15.1) | 2511 (18.2) | 3097 (20.4) | 2986 (22.6) | 722 (23.8) |

### Table S1

| Variable | Value | Total | March | April | May | June | July | August | September | October | November | December | 2021 |
|---|---|---|---|---|---|---|---|---|---|---|---|---|---|
| Seizures | Yes | 1821 (1.28) | 328 (1.21) | 544 (1.29) | 228 (1.85) | 96 (1.8) | 30 (1.07) | 25 (1.13) | 60 (1.14) | 164 (1.19) | 180 (1.19) | 146 (1.11) | 20 (0.659) |
| | No | 121559 (85.3) | 22794 (84.1) | 35061 (83) | 10408 (84.5) | 4709 (88.2) | 2606 (92.7) | 2031 (91.6) | 4735 (89.9) | 12203 (88.3) | 13136 (86.7) | 11271 (85.4) | 2605 (85.8) |
| | Unknown | 19160 (13.4) | 3986 (14.7) | 6662 (15.8) | 1675 (13.6) | 537 (10.1) | 175 (6.23) | 162 (7.3) | 470 (8.93) | 1455 (10.5) | 1839 (12.1) | 1788 (13.5) | 411 (13.5) |
| Shortness of breath | Yes | 92129 (64.6) | 17563 (64.8) | 28137 (66.6) | 7116 (57.8) | 3223 (60.3) | 1665 (59.2) | 1347 (60.7) | 3293 (62.5) | 8896 (64.4) | 9791 (64.6) | 8580 (65) | 2518 (82.9) |
| | No | 50146 (35.2) | 9469 (34.9) | 14087 (33.3) | 5178 (42.1) | 2116 (39.6) | 1142 (40.6) | 865 (39) | 1967 (37.4) | 4901 (35.5) | 5319 (35.1) | 4589 (34.8) | 513 (16.9) |
| | Unknown | 265 (0.186) | 76 (0.28) | 43 (0.102) | 17 (0.138) | 3 (0.0562) | 4 (0.142) | 6 (0.271) | 5 (0.095) | 25 (0.181) | 45 (0.297) | 36 (0.273) | 5 (0.165) |
| Ulcers | Yes | 2306 (1.62) | 396 (1.46) | 849 (2.01) | 317 (2.57) | 98 (1.83) | 25 (0.889) | 23 (1.04) | 44 (0.836) | 163 (1.18) | 186 (1.23) | 190 (1.44) | 15 (0.494) |
| | No | 105553 (74.1) | 18599 (68.6) | 31023 (73.4) | 8833 (71.7) | 3733 (69.9) | 1759 (62.6) | 1254 (56.5) | 4140 (78.6) | 11179 (80.9) | 12154 (80.2) | 10429 (79) | 2450 (80.7) |
| | Unknown | 34681 (24.3) | 8113 (29.9) | 10395 (24.6) | 3161 (25.7) | 1511 (28.3) | 1027 (36.5) | 941 (42.4) | 1081 (20.5) | 2480 (17.9) | 2815 (18.6) | 2586 (19.6) | 571 (18.8) |
| Vomiting | Yes | 25135 (17.6) | 4611 (17) | 7291 (17.2) | 2236 (18.2) | 893 (16.7) | 451 (16) | 376 (17) | 1130 (21.5) | 2589 (18.7) | 2750 (18.1) | 2250 (17) | 558 (18.4) |
| | No | 101164 (71) | 19063 (70.3) | 29401 (69.6) | 8676 (70.5) | 3985 (74.6) | 2212 (78.7) | 1705 (76.9) | 3764 (71.5) | 9988 (72.3) | 10829 (71.5) | 9425 (71.4) | 2116 (69.7) |
| | Unknown | 16241 (11.4) | 3434 (12.7) | 5575 (13.2) | 1399 (11.4) | 464 (8.69) | 148 (5.27) | 137 (6.18) | 371 (7.05) | 1245 (9.01) | 1576 (10.4) | 1530 (11.6) | 362 (11.9) |
| Wheezing | Yes | 9011 (6.32) | 2149 (7.93) | 2745 (6.49) | 718 (5.83) | 238 (4.46) | 120 (4.27) | 88 (3.97) | 313 (5.94) | 877 (6.34) | 929 (6.13) | 669 (5.07) | 165 (5.43) |
| | No | 109017 (76.5) | 19610 (72.3) | 31184 (73.8) | 9522 (77.3) | 4443 (83.2) | 2443 (86.9) | 1931 (87.1) | 4342 (82.5) | 11064 (80) | 11903 (78.5) | 10223 (77.4) | 2352 (77.5) |
| | Unknown | 24512 (17.2) | 5349 (19.7) | 8338 (19.7) | 2071 (16.8) | 661 (12.4) | 248 (8.82) | 199 (8.97) | 610 (11.6) | 1881 (13.6) | 2323 (15.3) | 2313 (17.5) | 519 (17.1) |

### Table S1

| Variable | Value | Total | March | April | May | June | July | August | September | October | November | December | 2021 |
|---|---|---|---|---|---|---|---|---|---|---|---|---|---|
| Asthma | Yes | 18147 (12.7) | 3621 (13.4) | 5129 (12.1) | 1369 (11.1) | 522 (9.77) | 266 (9.46) | 190 (8.57) | 642 (12.2) | 1924 (13.9) | 2166 (14.3) | 1827 (13.8) | 491 (16.2) |
| | No | 118096 (82.9) | 22265 (82.1) | 35126 (83.1) | 10366 (84.2) | 4621 (86.5) | 2477 (88.1) | 1949 (87.9) | 4432 (84.2) | 11439 (82.8) | 12338 (81.4) | 10672 (80.8) | 2411 (79.4) |
| | Unknown | 6297 (4.42) | 1222 (4.51) | 2012 (4.76) | 576 (4.68) | 199 (3.73) | 68 (2.42) | 79 (3.56) | 191 (3.63) | 459 (3.32) | 651 (4.3) | 706 (5.35) | 134 (4.41) |
| Chronic cardiac disease | Yes | 38747 (27.2) | 6870 (25.3) | 12110 (28.7) | 3891 (31.6) | 1361 (25.5) | 589 (21) | 406 (18.3) | 1265 (24) | 3849 (27.8) | 4220 (27.8) | 3614 (27.4) | 572 (18.8) |
| | No | 92500 (64.9) | 17327 (63.9) | 27209 (64.4) | 7447 (60.5) | 3510 (65.7) | 1859 (66.1) | 1502 (67.7) | 3551 (67.4) | 9163 (66.3) | 9949 (65.6) | 8682 (65.7) | 2301 (75.8) |
| | Unknown | 11293 (7.92) | 2911 (10.7) | 2948 (6.97) | 973 (7.9) | 471 (8.82) | 363 (12.9) | 310 (14) | 449 (8.53) | 810 (5.86) | 986 (6.51) | 909 (6.88) | 163 (5.37) |
| Chronic haemotologic disease | Yes | 5199 (3.65) | 988 (3.64) | 1636 (3.87) | 504 (4.09) | 187 (3.5) | 82 (2.92) | 40 (1.8) | 175 (3.32) | 449 (3.25) | 535 (3.53) | 521 (3.95) | 82 (2.7) |
| | No | 121364 (85.1) | 22156 (81.7) | 36418 (86.2) | 10174 (82.6) | 4357 (81.6) | 2116 (75.3) | 1616 (72.9) | 4449 (84.5) | 12299 (89) | 13420 (88.6) | 11617 (88) | 2742 (90.3) |
| | Unknown | 15977 (11.2) | 3964 (14.6) | 4213 (9.97) | 1633 (13.3) | 798 (14.9) | 613 (21.8) | 562 (25.3) | 641 (12.2) | 1074 (7.77) | 1200 (7.92) | 1067 (8.08) | 212 (6.98) |
| Chronic kidney disease | Yes | 21199 (14.9) | 3655 (13.5) | 6778 (16) | 2135 (17.3) | 760 (14.2) | 329 (11.7) | 221 (9.96) | 639 (12.1) | 2024 (14.6) | 2372 (15.7) | 1981 (15) | 305 (10) |
| | No | 114949 (80.6) | 22207 (81.9) | 33488 (79.2) | 9618 (78.1) | 4377 (81.9) | 2414 (85.9) | 1920 (86.6) | 4442 (84.4) | 11306 (81.8) | 12106 (79.9) | 10484 (79.4) | 2587 (85.2) |
| | Unknown | 6392 (4.48) | 1246 (4.6) | 2001 (4.73) | 558 (4.53) | 205 (3.84) | 68 (2.42) | 77 (3.47) | 184 (3.49) | 492 (3.56) | 677 (4.47) | 740 (5.6) | 144 (4.74) |
| Chronic neurological disorder | Yes | 15447 (10.8) | 2665 (9.83) | 5219 (12.3) | 1610 (13.1) | 496 (9.28) | 217 (7.72) | 131 (5.91) | 441 (8.38) | 1374 (9.94) | 1566 (10.3) | 1529 (11.6) | 199 (6.55) |
| | No | 120226 (84.3) | 23084 (85.2) | 34891 (82.5) | 10102 (82.1) | 4630 (86.7) | 2526 (89.9) | 2003 (90.3) | 4634 (88) | 11913 (86.2) | 12873 (84.9) | 10885 (82.4) | 2685 (88.4) |
| | Unknown | 6867 (4.82) | 1359 (5.01) | 2157 (5.1) | 599 (4.87) | 216 (4.04) | 68 (2.42) | 84 (3.79) | 190 (3.61) | 535 (3.87) | 716 (4.72) | 791 (5.99) | 152 (5.01) |
| Chronic pulmonary disease | Yes | 22434 (15.7) | 4171 (15.4) | 6734 (15.9) | 2098 (17) | 828 (15.5) | 332 (11.8) | 218 (9.83) | 769 (14.6) | 2374 (17.2) | 2554 (16.9) | 1962 (14.9) | 394 (13) |
| | No | 114135 (80.1) | 21823 (80.5) | 33624 (79.6) | 9681 (78.6) | 4319 (80.8) | 2408 (85.7) | 1922 (86.7) | 4309 (81.8) | 11021 (79.7) | 11992 (79.1) | 10523 (79.7) | 2513 (82.8) |
| | Unknown | 5971 (4.19) | 1114 (4.11) | 1909 (4.52) | 532 (4.32) | 195 (3.65) | 71 (2.53) | 78 (3.52) | 187 (3.55) | 427 (3.09) | 609 (4.02) | 720 (5.45) | 129 (4.25) |
| Dementia | Yes | 16538 (11.6) | 2488 (9.18) | 6367 (15.1) | 2068 (16.8) | 549 (10.3) | 196 (6.97) | 120 (5.41) | 384 (7.29) | 1336 (9.67) | 1507 (9.94) | 1389 (10.5) | 134 (4.41) |
| | No | 113546 (79.7) | 21434 (79.1) | 32616 (77.2) | 9153 (74.3) | 4283 (80.2) | 2257 (80.3) | 1783 (80.4) | 4433 (84.2) | 11569 (83.7) | 12496 (82.5) | 10811 (81.9) | 2711 (89.3) |
| | Unknown | 12456 (8.74) | 3186 (11.8) | 3284 (7.77) | 1090 (8.85) | 510 (9.55) | 358 (12.7) | 315 (14.2) | 448 (8.51) | 917 (6.63) | 1152 (7.6) | 1005 (7.61) | 191 (6.29) |
| Diabetes | Yes | 20417 (14.3) | 4679 (17.3) | 6350 (15) | 1585 (12.9) | 840 (15.7) | 454 (16.2) | 305 (13.8) | 737 (14) | 1730 (12.5) | 1806 (11.9) | 1551 (11.7) | 380 (12.5) |
| | No | 101400 (71.1) | 19193 (70.8) | 30540 (72.3) | 8583 (69.7) | 3299 (61.8) | 1431 (50.9) | 1054 (47.5) | 3673 (69.8) | 10287 (74.4) | 11222 (74) | 9836 (74.5) | 2282 (75.2) |
| | Unknown | 20723 (14.5) | 3236 (11.9) | 5377 (12.7) | 2143 (17.4) | 1203 (22.5) | 926 (32.9) | 859 (38.7) | 855 (16.2) | 1805 (13.1) | 2127 (14) | 1818 (13.8) | 374 (12.3) |
| HIV/AIDS | Yes | 525 (0.368) | 108 (0.398) | 169 (0.4) | 52 (0.422) | 16 (0.3) | 9 (0.32) | 10 (0.451) | 24 (0.456) | 45 (0.326) | 35 (0.231) | 47 (0.356) | 10 (0.329) |
| | No | 121496 (85.2) | 22284 (82.2) | 36330 (86) | 10262 (83.4) | 4191 (78.5) | 1875 (66.7) | 1347 (60.7) | 4450 (84.5) | 12380 (89.6) | 13703 (90.4) | 11898 (90.1) | 2776 (91.4) |
| | Unknown | 20519 (14.4) | 4716 (17.4) | 5768 (13.6) | 1997 (16.2) | 1135 (21.2) | 927 (33) | 861 (38.8) | 791 (15) | 1397 (10.1) | 1417 (9.35) | 1260 (9.54) | 250 (8.23) |
| Hypertension | Yes | 51415 (36.1) | 6233 (23) | 12665 (30) | 5487 (44.6) | 2408 (45.1) | 1226 (43.6) | 961 (43.3) | 2232 (42.4) | 6214 (45) | 6848 (45.2) | 5900 (44.7) | 1241 (40.9) |
| | No | 56904 (39.9) | 6744 (24.9) | 13465 (31.9) | 5880 (47.8) | 2628 (49.2) | 1441 (51.3) | 1169 (52.7) | 2740 (52) | 7014 (50.7) | 7586 (50.1) | 6592 (49.9) | 1645 (54.2) |
| | Unknown | 34221 (24) | 14131 (52.1) | 16137 (38.2) | 944 (7.67) | 306 (5.73) | 144 (5.12) | 88 (3.97) | 293 (5.57) | 594 (4.3) | 721 (4.76) | 713 (5.4) | 150 (4.94) |
| Liver disease | Yes | 4515 (3.17) | 827 (3.05) | 1267 (3) | 452 (3.67) | 174 (3.26) | 91 (3.24) | 64 (2.89) | 142 (2.7) | 455 (3.29) | 532 (3.51) | 439 (3.32) | 72 (2.37) |
| | No | 127065 (89.1) | 24099 (88.9) | 37799 (89.4) | 10694 (86.9) | 4656 (87.2) | 2377 (84.6) | 1831 (82.6) | 4734 (89.9) | 12569 (90.9) | 13718 (90.5) | 11813 (89.5) | 2775 (91.4) |
| | Unknown | 10960 (7.69) | 2182 (8.05) | 3201 (7.57) | 1165 (9.46) | 512 (9.58) | 343 (12.2) | 323 (14.6) | 389 (7.39) | 798 (5.77) | 905 (5.97) | 953 (7.22) | 189 (6.23) |
| Malignant neoplasm | Yes | 12548 (8.8) | 2453 (9.05) | 3736 (8.84) | 1220 (9.91) | 479 (8.97) | 201 (7.15) | 113 (5.09) | 350 (6.65) | 1132 (8.19) | 1441 (9.51) | 1218 (9.22) | 205 (6.75) |
| | No | 122870 (86.2) | 23260 (85.8) | 36281 (85.8) | 10426 (84.7) | 4637 (86.8) | 2539 (90.3) | 2016 (90.9) | 4709 (89.4) | 12160 (88) | 12977 (85.6) | 11189 (84.7) | 2676 (88.1) |
| | Unknown | 7122 (5) | 1395 (5.15) | 2250 (5.32) | 665 (5.4) | 226 (4.23) | 71 (2.53) | 89 (4.01) | 206 (3.91) | 530 (3.83) | 737 (4.86) | 798 (6.04) | 155 (5.11) |
| Malnutrition | Yes | 3125 (2.19) | 515 (1.9) | 1052 (2.49) | 341 (2.77) | 150 (2.81) | 42 (1.49) | 31 (1.4) | 94 (1.79) | 270 (1.95) | 297 (1.96) | 302 (2.29) | 31 (1.02) |
| | No | 119118 (83.6) | 22329 (82.4) | 35728 (84.5) | 10055 (81.7) | 4458 (83.5) | 2294 (81.6) | 1780 (80.3) | 4418 (83.9) | 11680 (84.5) | 12635 (83.4) | 11047 (83.7) | 2694 (88.7) |
| | Unknown | 20297 (14.2) | 4264 (15.7) | 5487 (13) | 1915 (15.6) | 734 (13.7) | 475 (16.9) | 407 (18.3) | 753 (14.3) | 1872 (13.5) | 2223 (14.7) | 1856 (14.1) | 311 (10.2) |
| Obesity | Yes | 17114 (12) | 3140 (11.6) | 4961 (11.7) | 1284 (10.4) | 582 (10.9) | 378 (13.4) | 247 (11.1) | 648 (12.3) | 1784 (12.9) | 1897 (12.5) | 1703 (12.9) | 490 (16.1) |
| | No | 100911 (70.8) | 18954 (69.9) | 30635 (72.5) | 8812 (71.6) | 3867 (72.4) | 1902 (67.7) | 1505 (67.9) | 3687 (70) | 9700 (70.2) | 10545 (69.6) | 9232 (69.9) | 2072 (68.2) |
| | Unknown | 24515 (17.2) | 5014 (18.5) | 6671 (15.8) | 2215 (18) | 893 (16.7) | 531 (18.9) | 466 (21) | 930 (17.7) | 2338 (16.9) | 2713 (17.9) | 2270 (17.2) | 474 (15.6) |
| Rheumatological disorder | Yes | 14118 (9.9) | 2285 (8.43) | 4091 (9.68) | 1260 (10.2) | 460 (8.61) | 169 (6.01) | 133 (6) | 462 (8.77) | 1594 (11.5) | 1763 (11.6) | 1597 (12.1) | 304 (10) |
| | No | 112159 (78.7) | 20793 (76.7) | 33822 (80) | 9430 (76.6) | 4087 (76.5) | 2030 (72.2) | 1521 (68.6) | 4161 (79) | 11131 (80.5) | 12155 (80.2) | 10509 (79.6) | 2520 (83) |
| | Unknown | 16263 (11.4) | 4030 (14.9) | 4354 (10.3) | 1621 (13.2) | 795 (14.9) | 612 (21.8) | 564 (25.4) | 642 (12.2) | 1097 (7.94) | 1237 (8.16) | 1099 (8.32) | 212 (6.98) |
| Smoking | Yes | 7401 (5.19) | 1409 (5.2) | 2192 (5.19) | 749 (6.08) | 326 (6.1) | 156 (5.55) | 138 (6.22) | 256 (4.86) | 660 (4.77) | 726 (4.79) | 687 (5.2) | 102 (3.36) |
| | No | 50930 (35.7) | 12805 (47.2) | 15940 (37.7) | 3592 (29.2) | 1730 (32.4) | 1082 (38.5) | 953 (43) | 1881 (35.7) | 3929 (28.4) | 4240 (28) | 3713 (28.1) | 1065 (35.1) |
| | Unknown | 84209 (59.1) | 12894 (47.6) | 24135 (57.1) | 7970 (64.7) | 3286 (61.5) | 1573 (56) | 1127 (50.8) | 3128 (59.4) | 9233 (66.8) | 10189 (67.2) | 8805 (66.7) | 1869 (61.6) |

TABLE S1

| Variable | Value | Total | March | April | May | June | July | August | September | October | November | December | 2021 |
|---|---|---|---|---|---|---|---|---|---|---|---|---|---|
| Country | Argentina | 54 (0.0379) | 1 (0.00369) | 2 (0.00473) | 3 (0.0244) | 6 (0.112) | 6 (0.213) | 13 (0.586) | 13 (0.247) | 10 (0.0723) | 0 (0) | 0 (0) | 0 (0) |
| | Austria | 26 (0.0182) | 5 (0.0184) | 3 (0.0071) | 3 (0.0244) | 0 (0) | 1 (0.0356) | 3 (0.135) | 3 (0.057) | 6 (0.0434) | 2 (0.0132) | 0 (0) | 0 (0) |
| | Belgium | 1201 (0.843) | 346 (1.28) | 396 (0.937) | 79 (0.642) | 3 (0.0562) | 6 (0.213) | 18 (0.812) | 23 (0.437) | 198 (1.43) | 115 (0.759) | 17 (0.129) | 0 (0) |
| | Brazil | 600 (0.421) | 93 (0.343) | 177 (0.419) | 203 (1.65) | 76 (1.42) | 27 (0.961) | 20 (0.902) | 3 (0.057) | 1 (0.00723) | 0 (0) | 0 (0) | 0 (0) |
| | Canada | 3128 (2.19) | 419 (1.55) | 1065 (2.52) | 385 (3.13) | 76 (1.42) | 78 (2.77) | 43 (1.94) | 169 (3.21) | 316 (2.29) | 340 (2.24) | 202 (1.53) | 35 (1.15) |
| | Chile | 132 (0.0926) | 6 (0.0221) | 11 (0.026) | 90 (0.731) | 25 (0.468) | 0 (0) | 0 (0) | 0 (0) | 0 (0) | 0 (0) | 0 (0) | 0 (0) |
| | Colombia | 436 (0.306) | 21 (0.0775) | 49 (0.116) | 43 (0.349) | 111 (2.08) | 84 (2.99) | 44 (1.98) | 14 (0.266) | 34 (0.246) | 20 (0.132) | 11 (0.0833) | 5 (0.165) |
| | Czechia | 20 (0.014) | 0 (0) | 0 (0) | 2 (0.0162) | 4 (0.0749) | 11 (0.391) | 3 (0.135) | 0 (0) | 0 (0) | 0 (0) | 0 (0) | 0 (0) |
| | Dominican Republic | 13 (0.00912) | 4 (0.0148) | 9 (0.0213) | 0 (0) | 0 (0) | 0 (0) | 0 (0) | 0 (0) | 0 (0) | 0 (0) | 0 (0) | 0 (0) |
| | Ecuador | 27 (0.0189) | 3 (0.0111) | 23 (0.0544) | 0 (0) | 0 (0) | 0 (0) | 0 (0) | 0 (0) | 1 (0.00723) | 0 (0) | 0 (0) | 0 (0) |
| | Estonia | 62 (0.0435) | 16 (0.059) | 13 (0.0308) | 2 (0.0162) | 1 (0.0187) | 0 (0) | 0 (0) | 2 (0.038) | 4 (0.0289) | 8 (0.0528) | 7 (0.053) | 9 (0.296) |
| | France | 2965 (2.08) | 1935 (7.14) | 880 (2.08) | 51 (0.414) | 20 (0.374) | 18 (0.64) | 25 (1.13) | 36 (0.684) | 0 (0) | 0 (0) | 0 (0) | 0 (0) |
| | Germany | 128 (0.0898) | 39 (0.144) | 44 (0.104) | 13 (0.106) | 6 (0.112) | 0 (0) | 3 (0.135) | 5 (0.095) | 8 (0.0579) | 8 (0.0528) | 2 (0.0151) | 0 (0) |
| | Ghana | 2 (0.0014) | 0 (0) | 0 (0) | 0 (0) | 2 (0.0374) | 0 (0) | 0 (0) | 0 (0) | 0 (0) | 0 (0) | 0 (0) | 0 (0) |
| | Greece | 40 (0.0281) | 8 (0.0295) | 21 (0.0497) | 10 (0.0812) | 1 (0.0187) | 0 (0) | 0 (0) | 0 (0) | 0 (0) | 0 (0) | 0 (0) | 0 (0) |
| | Hong Kong | 24 (0.0168) | 16 (0.059) | 4 (0.00946) | 0 (0) | 0 (0) | 4 (0.142) | 0 (0) | 0 (0) | 0 (0) | 0 (0) | 0 (0) | 0 (0) |
| | India | 1757 (1.23) | 5 (0.0184) | 11 (0.026) | 48 (0.39) | 261 (4.89) | 304 (10.8) | 460 (20.7) | 303 (5.75) | 169 (1.22) | 117 (0.772) | 77 (0.583) | 2 (0.0659) |
| | Indonesia | 505 (0.354) | 68 (0.251) | 73 (0.173) | 34 (0.276) | 31 (0.58) | 50 (1.78) | 90 (4.06) | 62 (1.18) | 45 (0.326) | 15 (0.099) | 32 (0.242) | 5 (0.165) |
| | Ireland | 1063 (0.746) | 360 (1.33) | 422 (0.998) | 86 (0.699) | 18 (0.337) | 6 (0.213) | 9 (0.406) | 43 (0.817) | 64 (0.463) | 46 (0.304) | 5 (0.0379) | 4 (0.132) |
| | Israel | 401 (0.281) | 55 (0.203) | 94 (0.222) | 16 (0.13) | 43 (0.805) | 146 (5.19) | 47 (2.12) | 0 (0) | 0 (0) | 0 (0) | 0 (0) | 0 (0) |
| | Italy | 558 (0.391) | 344 (1.27) | 89 (0.211) | 7 (0.0569) | 2 (0.0374) | 4 (0.142) | 2 (0.0902) | 13 (0.247) | 40 (0.289) | 38 (0.251) | 16 (0.121) | 3 (0.0988) |
| | Japan | 88 (0.0617) | 23 (0.0848) | 31 (0.0733) | 7 (0.0569) | 7 (0.131) | 9 (0.32) | 6 (0.271) | 1 (0.019) | 0 (0) | 4 (0.0264) | 0 (0) | 0 (0) |
| | Kuwait | 39 (0.0274) | 10 (0.0369) | 15 (0.0355) | 2 (0.0162) | 0 (0) | 1 (0.0356) | 0 (0) | 5 (0.095) | 1 (0.00723) | 1 (0.0066) | 4 (0.0303) | 0 (0) |
| | Malaysia | 2 (0.0014) | 0 (0) | 1 (0.00237) | 0 (0) | 0 (0) | 0 (0) | 0 (0) | 0 (0) | 0 (0) | 1 (0.0066) | 0 (0) | 0 (0) |
| | Mexico | 12 (0.00842) | 1 (0.00369) | 4 (0.00946) | 4 (0.0325) | 2 (0.0374) | 0 (0) | 0 (0) | 0 (0) | 0 (0) | 1 (0.0066) | 0 (0) | 0 (0) |
| | Nepal | 605 (0.424) | 0 (0) | 0 (0) | 0 (0) | 4 (0.0749) | 17 (0.605) | 59 (2.66) | 90 (1.71) | 154 (1.11) | 177 (1.17) | 104 (0.788) | 0 (0) |
| | Netherlands | 1823 (1.28) | 846 (3.12) | 870 (2.06) | 71 (0.577) | 5 (0.0936) | 7 (0.249) | 7 (0.316) | 5 (0.095) | 12 (0.0868) | 0 (0) | 0 (0) | 0 (0) |
| | New Zealand | 20 (0.014) | 2 (0.00738) | 7 (0.0166) | 0 (0) | 1 (0.0187) | 0 (0) | 6 (0.271) | 4 (0.076) | 0 (0) | 0 (0) | 0 (0) | 0 (0) |
| | Norway | 343 (0.241) | 129 (0.476) | 90 (0.213) | 11 (0.0894) | 7 (0.131) | 2 (0.0711) | 5 (0.225) | 13 (0.247) | 14 (0.101) | 43 (0.284) | 26 (0.197) | 3 (0.0988) |
| | Pakistan | 2066 (1.45) | 6 (0.0221) | 59 (0.14) | 285 (2.32) | 547 (10.2) | 253 (9) | 89 (4.01) | 82 (1.56) | 96 (0.695) | 278 (1.83) | 335 (2.54) | 36 (1.19) |
| | Peru | 290 (0.203) | 7 (0.0258) | 88 (0.208) | 138 (1.12) | 21 (0.393) | 19 (0.676) | 15 (0.676) | 0 (0) | 2 (0.0145) | 0 (0) | 0 (0) | 0 (0) |
| | Poland | 283 (0.199) | 213 (0.786) | 55 (0.13) | 1 (0.00812) | 0 (0) | 4 (0.142) | 8 (0.361) | 1 (0.019) | 1 (0.00723) | 0 (0) | 0 (0) | 0 (0) |
| | Portugal | 746 (0.523) | 251 (0.926) | 173 (0.409) | 80 (0.65) | 78 (1.46) | 50 (1.78) | 36 (1.62) | 29 (0.551) | 41 (0.297) | 7 (0.0462) | 0 (0) | 1 (0.0329) |
| | Qatar | 154 (0.108) | 33 (0.122) | 117 (0.277) | 4 (0.0325) | 0 (0) | 0 (0) | 0 (0) | 0 (0) | 0 (0) | 0 (0) | 0 (0) | 0 (0) |
| | Romania | 861 (0.604) | 103 (0.38) | 193 (0.457) | 139 (1.13) | 143 (2.68) | 167 (5.94) | 90 (4.06) | 22 (0.418) | 3 (0.0217) | 1 (0.0066) | 0 (0) | 0 (0) |
| | Saudi Arabia | 149 (0.105) | 0 (0) | 4 (0.00946) | 62 (0.504) | 82 (1.54) | 1 (0.0356) | 0 (0) | 0 (0) | 0 (0) | 0 (0) | 0 (0) | 0 (0) |
| | South Africa | 196 (0.138) | 0 (0) | 8 (0.0189) | 55 (0.447) | 72 (1.35) | 42 (1.49) | 18 (0.812) | 1 (0.019) | 0 (0) | 0 (0) | 0 (0) | 0 (0) |
| | South Korea | 39 (0.0274) | 13 (0.048) | 3 (0.0071) | 0 (0) | 3 (0.0562) | 7 (0.249) | 4 (0.18) | 7 (0.133) | 1 (0.00723) | 1 (0.0066) | 0 (0) | 0 (0) |
| | Spain | 400 (0.281) | 341 (1.26) | 24 (0.0568) | 2 (0.0162) | 2 (0.0374) | 3 (0.107) | 2 (0.0902) | 7 (0.133) | 11 (0.0796) | 7 (0.0462) | 1 (0.00757) | 0 (0) |
| | Taiwan | 1 (0.000702) | 0 (0) | 1 (0.00237) | 0 (0) | 0 (0) | 0 (0) | 0 (0) | 0 (0) | 0 (0) | 0 (0) | 0 (0) | 0 (0) |
| | Thailand | 8 (0.00561) | 6 (0.0221) | 0 (0) | 0 (0) | 0 (0) | 0 (0) | 0 (0) | 1 (0.019) | 0 (0) | 0 (0) | 1 (0.00757) | 0 (0) |
| | Turkey | 1 (0.000702) | 1 (0.00369) | 0 (0) | 0 (0) | 0 (0) | 0 (0) | 0 (0) | 0 (0) | 0 (0) | 0 (0) | 0 (0) | 0 (0) |
| | United Kingdom | 118137 (82.9) | 19466 (71.8) | 36549 (86.5) | 10176 (82.7) | 3586 (67.1) | 1389 (49.4) | 1028 (46.3) | 4266 (81) | 12544 (90.8) | 13881 (91.6) | 12321 (93.3) | 2931 (96.5) |
| | Ukraine | 84 (0.0589) | 0 (0) | 0 (0) | 0 (0) | 0 (0) | 0 (0) | 0 (0) | 0 (0) | 6 (0.0434) | 35 (0.231) | 42 (0.318) | 1 (0.0329) |
| | United States of America | 3050 (2.14) | 1912 (7.05) | 589 (1.39) | 199 (1.62) | 96 (1.8) | 95 (3.38) | 65 (2.93) | 42 (0.798) | 40 (0.289) | 9 (0.0594) | 2 (0.0151) | 1 (0.0329) |
| | Viet Nam | 1 (0.000702) | 1 (0.00369) | 0 (0) | 0 (0) | 0 (0) | 0 (0) | 0 (0) | 0 (0) | 0 (0) | 0 (0) | 0 (0) | 0 (0) |
